# Population-Scale, Genotype-First Characterization of Monogenic Diabetes in 374,973 Multi-Ancestry Individuals from the *All of Us* Research Program

**DOI:** 10.64898/2026.06.12.26355541

**Authors:** Masashi Hasebe, Satoshi Yoshiji

## Abstract

**OBJECTIVE:** To characterize the prevalence and penetrance of maturity-onset diabetes of the young (MODY) in a multi-ancestry population using a genotype-first design.

**RESEARCH DESIGN AND METHODS:** We analyzed whole-genome sequencing and clinical data from 374,973 unrelated *All of Us* participants (42.0% non-European ancestry). We identified pathogenic or likely pathogenic (P/LP) variants in 10 established MODY genes and assessed carrier prevalence, diabetes penetrance, and glycemic profiles. We evaluated age-dependent diabetes risk by comparing carriers with non-carriers stratified by type 2 diabetes polygenic risk score (T2D PRS).

**RESULTS:** We identified 370 carriers of P/LP MODY gene variants (0.099%; 1 in 1,013), with similar carrier prevalence among European- and African-ancestry participants (0.105% in both groups). Diabetes penetrance was incomplete (13.4% by age 40; 43.5% by age 60) and varied by etiology: highest for *GCK* (56.0% by age 60), intermediate for HNF genes (*HNF1A*/*HNF1B*/*HNF4A*; 45.4%), and lowest for non-GCK/HNF genes (*ABCC8*/*INS*/*KCNJ11*/*NEUROD1*/*PDX1*/*RFX6*; 29.0%). In multivariable Cox models using non-carriers in the middle 80% of the T2D PRS as the reference, non-GCK/HNF gene variant carriers had modestly increased diabetes risk (HR, 1.57), similar to non-carriers in the top 10% of T2D PRS (HR, 1.64). These associations were observed in both European- and non-European-ancestry individuals. HbA1c profiles differed by etiology, with stable mild hyperglycemia in *GCK* variant carriers and greater variability among HNF and non-GCK/HNF gene variant carriers.

**CONCLUSIONS:** MODY gene variants showed incomplete, etiology-dependent penetrance across ancestries. Carriers of P/LP variants in lower-penetrance genes had diabetes risk comparable to that of non-carriers with high polygenic susceptibility.

**Graphical Abstract:** 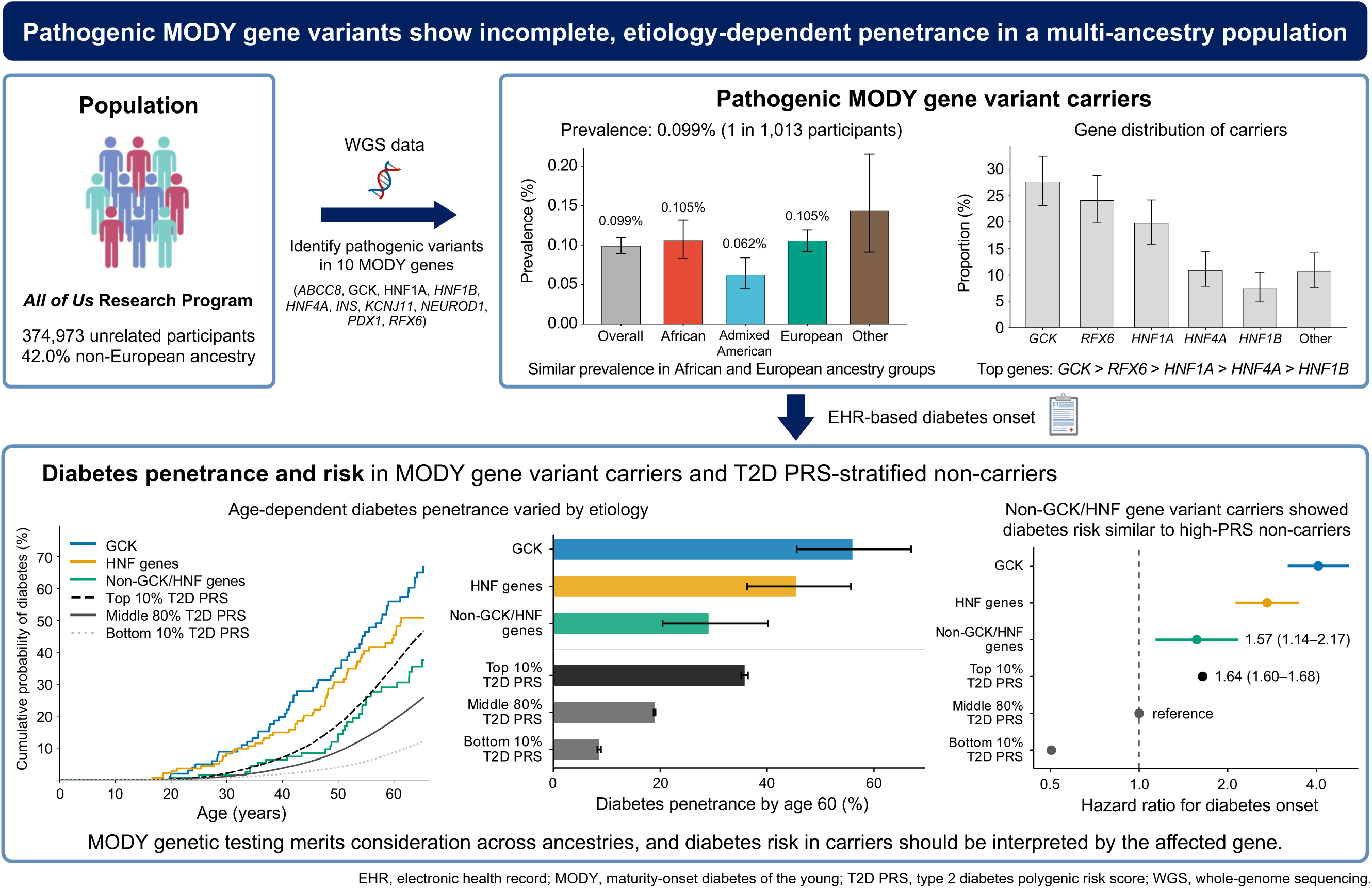

## INTRODUCTION

Monogenic diabetes comprises forms of diabetes primarily caused by pathogenic variants in single genes. Maturity-onset diabetes of the young (MODY), the most common form of monogenic diabetes, is usually inherited in an autosomal dominant pattern and has been attributed to pathogenic variants in multiple established genes (1). Because MODY has clinical implications that differ from those of common multifactorial diabetes, a genetic diagnosis of MODY can refine diabetes classification and guide treatment, prognosis, and cascade testing for relatives. Historically, most MODY studies have followed a phenotype-first design, in which genetic testing follows clinical suspicion of monogenic diabetes, particularly early-onset diabetes or a strong family history (2). These clinically selected studies have reported diagnostic yields of approximately 20–30% and have shaped current understanding of MODY prevalence, penetrance, and clinical spectrum (3, 4).

In rare genetic diseases, phenotype-first ascertainment tends to include individuals whose presentation is suggestive enough to prompt genetic testing. This selection can overestimate penetrance and underrepresent mildly affected or unaffected carriers (5). Population-based genotype-first studies address this limitation by identifying variant carriers independently of the presenting phenotype, allowing penetrance and expressivity to be assessed with less influence from clinical ascertainment (6, 7). Reflecting this difference in ascertainment, genotype-first studies of MODY genes have shown lower and more heterogeneous diabetes penetrance than phenotype-first studies (5, 8–11).

Although large-scale genotype-first studies of MODY genes have provided valuable insights, many have been conducted in predominantly European-ancestry cohorts, including the UK Biobank (5, 9–11). Consequently, the prevalence and phenotypic spectrum of pathogenic MODY gene variants remain incompletely defined in population-scale cohorts with greater ancestral diversity (12). Recent rare-variant studies have further highlighted the importance of studying monogenic diabetes genes in diverse populations, because some rare, large-effect variants may be population-specific or enriched in ancestry groups that remain underrepresented in genetic studies (13). Phenotype-first studies from non-European populations also suggest variation across populations in the proportion of diabetes cases carrying pathogenic variants in MODY genes, as well as heterogeneity in carrier phenotypes (14–17). These findings highlight the need for population-scale genotype-first studies in multi-ancestry cohorts to define the prevalence and carrier phenotypes of pathogenic MODY gene variants beyond predominantly European-ancestry populations.

Here, we conducted a genotype-first study of 10 established MODY genes using genetic and clinical data from 374,973 unrelated participants in the multi-ancestry *All of Us* Research Program. We aimed to identify pathogenic or likely pathogenic (P/LP) variants and assess carrier prevalence, age-dependent diabetes risk, and glycemic phenotypes in a diverse U.S. population. We further placed these phenotypic features in a population-wide context by comparing carriers with non-carriers stratified by common-variant polygenic susceptibility to type 2 diabetes.

## RESEARCH DESIGN AND METHODS

### Study Participants

This study used Controlled Tier Curated Data Repository version 8 from the *All of Us* Research Program, a large and diverse U.S. research cohort launched by the National Institutes of Health in 2018 (18). We included adult participants with linked electronic health record (EHR) data and available short-read whole-genome sequencing (srWGS) data from blood-derived DNA generated on the Illumina NovaSeq 6000 platform under standardized procedures (19). We further required available genetic principal components (PCs) and genetic ancestry assignments provided by *All of Us*, excluded participants flagged for low-quality srWGS data, and removed closely related individuals using the *All of Us* relatedness table (kinship score >0.1). Genetic ancestry was assigned using the *All of Us* auxiliary genomic ancestry classifications, based on genetic similarity to global reference populations, and grouped as African, Admixed American, European, South Asian, East Asian, or Middle Eastern. The final study population included 374,973 unrelated participants.

The *All of Us* Research Program was approved by the *All of Us* Institutional Review Board. All participants provided informed consent, including authorization for research use of EHR and other external health data.

### MODY Gene Variant Extraction and Classification

We evaluated single-nucleotide variants (SNVs) and indels in 10 established MODY genes: *ABCC8*, *GCK*, *HNF1A*, *HNF1B*, *HNF4A*, *INS*, *KCNJ11*, *NEUROD1*, *PDX1*, and *RFX6* (1). *CEL* was not included because pathogenic *CEL*-MODY variants typically occur in the variable number tandem repeat region, which is not reliably assessed using standard srWGS data (20). Candidate variants in the 10 target genes were first screened using the *All of Us* Variant Annotation Table (VAT) (21). The VAT provides functional annotations for passing SNVs and indels from the *All of Us* srWGS data. Included variants must pass site-level filters and genotype-level filter tags, and sites with 50 or more alternate alleles are excluded. The initial candidate set was restricted to VAT-represented passing SNVs and indels to minimize technically unreliable variant calls.

From the VAT records, we extracted variants in the 10 target genes that overlapped coding regions or canonical splice donor or acceptor sites. Variant consequences were annotated using Ensembl Variant Effect Predictor with the GRCh38 cache release 115 (22) and the Loss-Of-Function Transcript Effect Estimator plugin (LOFTEE) (23). ClinVar assertions were updated by exact matching on chromosome, position, reference allele, and alternate allele against the GRCh38 ClinVar VCF downloaded on May 7, 2026 (24).

To limit false positives, we restricted the candidate set to predicted protein-truncating or canonical splice donor or acceptor variants with LOFTEE high-confidence annotations and ultra-rare missense variants with a maximum gnomAD population allele frequency <0.001% based on gnomAD allele-frequency fields in the VAT. Variants retained after screening were manually reviewed according to the ACMG/AMP guidelines (25, 26), with applicable ClinGen guidance (27) and available variant-level evidence. Details of the classification approach are provided in **Supplementary Note 1**. Phenotypes of *All of Us* participants were not used for variant classification.

Participants carrying P/LP sequence variants were then identified from the *All of Us* srWGS genotype files by matching the corresponding alternate allele. After genotype-level and sequencing-read quality control, carriers who met the main study inclusion criteria were included in the analysis. Details of genotype-level and sequencing-read quality control are provided in **Supplementary Note 2**.

In addition to SNVs and indels, we evaluated 17q12 deletions because they account for a substantial proportion of *HNF1B*-MODY (28). Among participants with available srWGS structural-variant data, those with a heterozygous deletion spanning *HNF1B* at 17q12 were included as carriers of pathogenic *HNF1B* variants. Details of 17q12 deletion ascertainment are provided in **Supplementary Note 3**.

### Calculation of Type 2 Diabetes Polygenic Risk Score

We calculated a type 2 diabetes polygenic risk score (T2D PRS) using multi-ancestry allelic weights for 1,289 independent genome-wide significant index variants from the largest multi-ancestry genome-wide association study meta-analysis of type 2 diabetes, which did not include *All of Us* participants (29). Variants were matched to the *All of Us* srWGS data by GRCh38 chromosome, position, and allele orientation. Of the 1,289 variants, 1,284 were available and allele-matched in *All of Us*. The T2D PRS was calculated in PLINK v2.0 as a weighted sum of effect-allele dosages (30).

### Diabetes Phenotype Definitions

Under the primary definition, diabetes was defined by any of the following criteria: a type 2 diabetes diagnosis code in the EHR, self-reported type 2 diabetes, glycated hemoglobin (HbA1c) ≥6.5%, fasting glucose ≥126 mg/dL, or non-insulin glucose-lowering medication use. The earliest qualifying record defined age at diabetes ascertainment. Diagnosis codes and self-reported history were restricted to type 2 diabetes because MODY is often clinically labeled as type 2 diabetes in routine care (31). Insulin use alone and random or otherwise unspecified glucose values ≥200 mg/dL were not used to define first recorded diabetes, because these records can reflect acute-care hyperglycemia or its treatment and do not reliably indicate chronic diabetes when used alone.

We used two alternative diabetes definitions. One excluded non-insulin glucose-lowering medication use, because some of these medications may be prescribed for other indications. The other used diagnosis codes and self-reported diabetes records without restricting diabetes subtype, to account for possible documentation under other diabetes labels. Full details are provided in **Supplementary Note 4**.

### Statistical Analysis

Carrier prevalence was calculated overall and by sex and ancestry group as the proportion of participants carrying a P/LP variant in any evaluated MODY gene. Exact binomial 95% confidence intervals (CIs) were calculated for prevalence and gene-distribution proportions. Differences by sex were evaluated using Fisher’s exact test, and variation in carrier prevalence across ancestry groups was evaluated using a chi-square test. To comply with the *All of Us* Data and Statistics Dissemination Policy, results for a reported category with fewer than 20 participants were not shown or were combined into broader categories.

Distinct P/LP variants were summarized descriptively, including sharing across ancestry groups and distributions by gene group and variant type. For variant summaries and phenotype analyses, genes were grouped as *GCK*, HNF genes (*HNF1A*, *HNF1B*, and *HNF4A*), and non-GCK/HNF genes (*ABCC8*, *INS*, *KCNJ11*, *NEUROD1*, *PDX1*, and *RFX6*). This grouping reflected lifelong mild hyperglycemia in *GCK*-MODY, relatively high diabetes penetrance in HNF-related MODY, and rarity or heterogeneous penetrance among the other genes (9, 10, 32, 33).

For analyses comparing MODY gene variant carriers with non-carriers, non-carriers were stratified into the bottom 10%, middle 80% (hereafter, the middle group), and top 10% of the T2D PRS distribution among non-carriers. In regression models, non-carriers in the middle group were used as the reference.

Diabetes penetrance was estimated using Kaplan-Meier methods for the primary and two alternative diabetes definitions. Kaplan-Meier curves and log-rank tests were generated for the three carrier groups and the three non-carrier groups defined by T2D PRS. In accordance with the *All of Us* Data and Statistics Dissemination Policy, curves were truncated before any displayed group had fewer than 20 participants at risk. Diabetes penetrance at ages 40 and 60 was reported for all MODY gene variant carriers and non-carriers stratified by T2D PRS. For the three carrier gene groups, only penetrance at age 60 was reported because estimates at age 40 were based on sparse events. We also calculated the proportion of participants with diabetes by ages 40 and 60 who carried a P/LP variant in any evaluated MODY gene, using all participants with diabetes by each age threshold as the denominator. Exact binomial 95% CIs were calculated for these proportions.

Age-dependent diabetes onset, based on age at diabetes ascertainment, was further evaluated using Cox proportional hazards models with age as the time scale. Cox models were adjusted for sex and the first 10 genetic PCs. In ancestry-stratified analyses, the same models were fitted separately among participants with European ancestry and those not assigned to European ancestry. Delayed-entry Cox models were used as a sensitivity analysis, with entry at the start of linked EHR follow-up and exit at the earliest qualifying diabetes record during follow-up or at the end of follow-up. The proportional hazards assumption was assessed by visual inspection of Schoenfeld residuals.

Glycemic profiles were evaluated among participants with at least three HbA1c measurements separated by at least 90 days. For each participant, we calculated median HbA1c, maximum HbA1c, and within-person HbA1c standard deviation (SD). For each HbA1c measure, adjusted mean differences were estimated using linear regression models with non-carriers in the middle group as the reference; models included age at the midpoint of the HbA1c observation window, sex, and the first 10 genetic PCs, with robust HC3 standard errors.

All *P* values were two-sided. A *P* value <0.05 was used as the threshold for statistical significance. Analyses were performed using Python version 3.10.16. This study followed the STrengthening the REporting of Genetic Association Studies (STREGA) guidelines (34).

### Data and Resource Availability

All analyses used de-identified participant data accessed through the *All of Us* Researcher Workbench, Controlled Tier, version 8, in accordance with program policies.

## RESULTS

### Prevalence and Distribution of P/LP Variants in MODY Genes

Among 374,973 unrelated *All of Us* participants, the median age at recruitment was 53.0 years, 60.2% were female, and 42.0% were assigned to non-European ancestry groups (**Supplementary Table 1**). We identified 203 distinct P/LP variants in the evaluated MODY genes: 202 SNVs or indels and the recurrent 17q12 deletion involving *HNF1B* (**Supplementary Table 2**). Overall, 370 participants carried one of these variants, corresponding to a prevalence of 0.099% (95% CI, 0.089 to 0.109), or approximately 1 in 1,013 participants. Among carriers, 142 (38.4%) were assigned to a non-European ancestry group. Carrier prevalence was similar among male and female participants: 0.103% and 0.096%, respectively (*P* = 0.56) (**Supplementary Table 3**).

By ancestry group, carrier prevalence was 0.105% among African, 0.062% among Admixed American, 0.105% among European, and 0.143% in the combined other-ancestry group (*P* for heterogeneity = 0.0034) (**Figure 1A** and **Supplementary Table 3**). *GCK* accounted for the largest proportion of carriers (*n* = 102; 27.6%), followed by *RFX6* (*n* = 89; 24.1%), *HNF1A* (*n* = 73; 19.7%), *HNF4A* (*n* = 40; 10.8%), *HNF1B* (*n* = 27; 7.3%), and the remaining five genes combined (*n* = 39; 10.5%) (**Figure 1B** and **Supplementary Table 4**).

**Figure 1.**
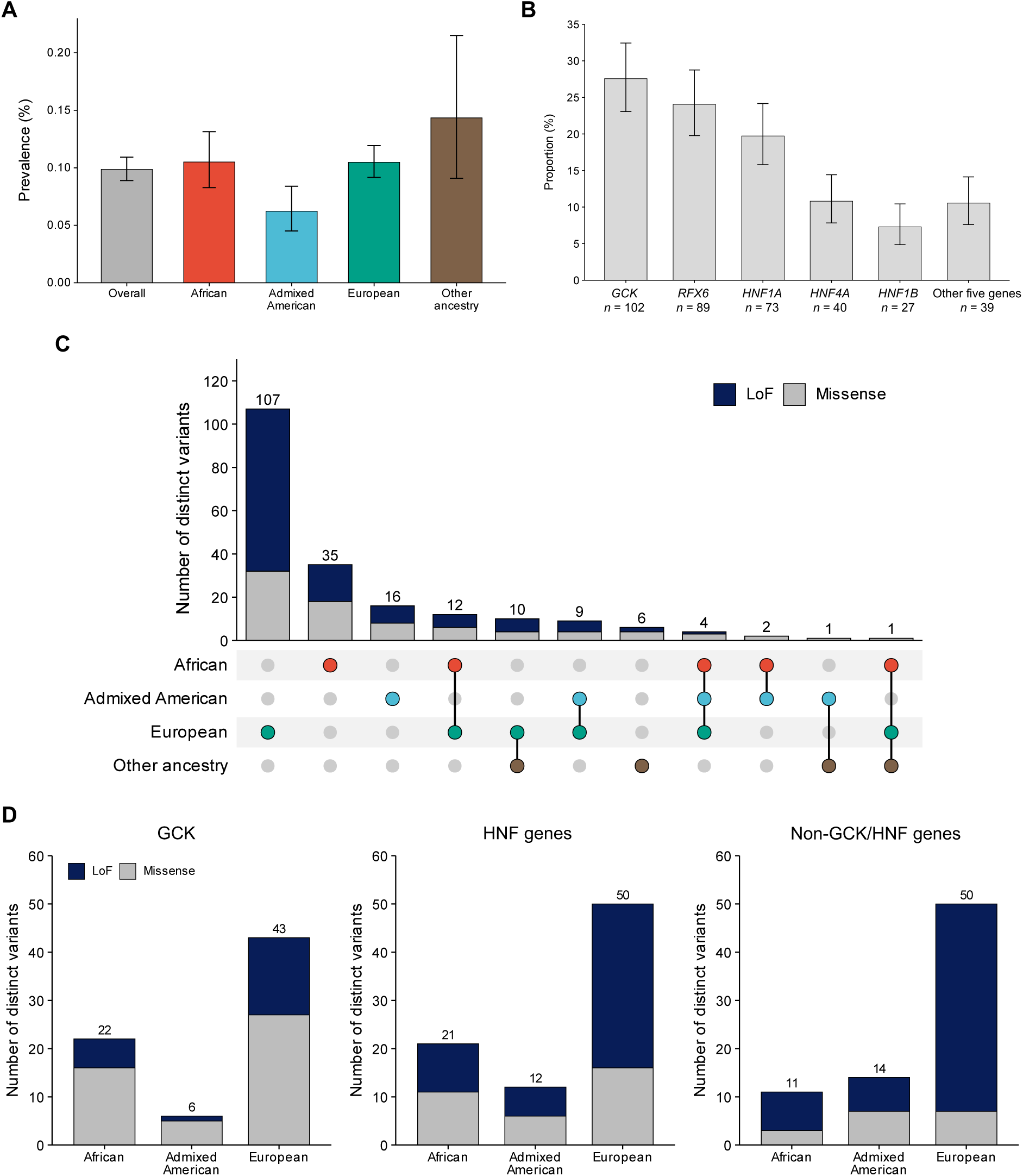
Prevalence and distribution of pathogenic or likely pathogenic variants in MODY genes among *All of Us* participants. Carrier prevalence for pathogenic or likely pathogenic (P/LP) variants in MODY genes is shown overall (370/374,973) and by ancestry (76/72,346 African; 43/69,006 Admixed American; 228/217,586 European; 23/16,035 other ancestry; *P* = 0.0034 for heterogeneity across ancestry groups) (A). The distribution of carriers by gene or gene category is shown as the proportion among all 370 carriers (B). The UpSet plot shows the overlap of distinct P/LP variants across ancestry categories; vertical bars indicate the number of distinct variants observed in each ancestry combination, stratified by variant type, and filled dots identify the ancestries represented in that combination (C). Distinct P/LP variant counts are shown by ancestry, gene category, and variant type for African, Admixed American, and European ancestry categories; the combined other ancestry category is not shown in this detailed breakdown because of sparse data (D). In panels A and B, bars indicate proportions and error bars indicate 95% confidence intervals (CIs). In panels C and D, counts represent distinct variants, not carrier counts, and should not be interpreted as ancestry-by-gene participant counts. The recurrent 17q12 deletion involving *HNF1B* was counted as one distinct variant and included in the loss-of-function (LoF) category. HNF genes include *HNF1A*, *HNF1B*, and *HNF4A*. Non-GCK/HNF genes include *ABCC8*, *INS*, *KCNJ11*, *NEUROD1*, *PDX1*, and *RFX6*.

Most distinct P/LP variants were observed in only one ancestry category (164/203; 80.8%): 107 among European, 35 among African, 16 among Admixed American, and 6 among participants in other ancestry groups (**Figure 1C**). Overall, 60 distinct variants (29.6%) were observed only among participants with non-European ancestry. Across ancestry categories, *GCK* variants were mainly missense, whereas loss-of-function variants accounted for a larger proportion of variants in HNF and non-GCK/HNF categories; in the latter, this was driven largely by *RFX6* (**Figure 1D** and **Supplementary Table 2**).

### Age-dependent Diabetes Penetrance

Using the primary definition, Kaplan-Meier curves showed the highest diabetes penetrance for *GCK* variant carriers, followed by HNF gene variant carriers; non-GCK/HNF gene variant carriers showed a more modest increase, closer to that of non-carriers in the top 10% of T2D PRS than to the other carrier groups (**Figure 2A**). Among all MODY gene variant carriers, penetrance was 13.4% by age 40 and 43.5% by age 60; by comparison, penetrance among non-carriers in the top 10% of T2D PRS was 7.1% and 35.8%, respectively (**Supplementary Figure 1** and **Supplementary Table 5**). Within carriers, penetrance by age 60 differed by genetic etiology: 56.0% for *GCK*, 45.4% for HNF genes, and 29.0% for non-GCK/HNF genes (**Figure 2B** and **Supplementary Table 5**). Sensitivity analyses using the medication-excluded and broader diabetes-label definitions showed similar patterns (**Supplementary Figure 2** and **Supplementary Table 5**).

**Figure 2.**
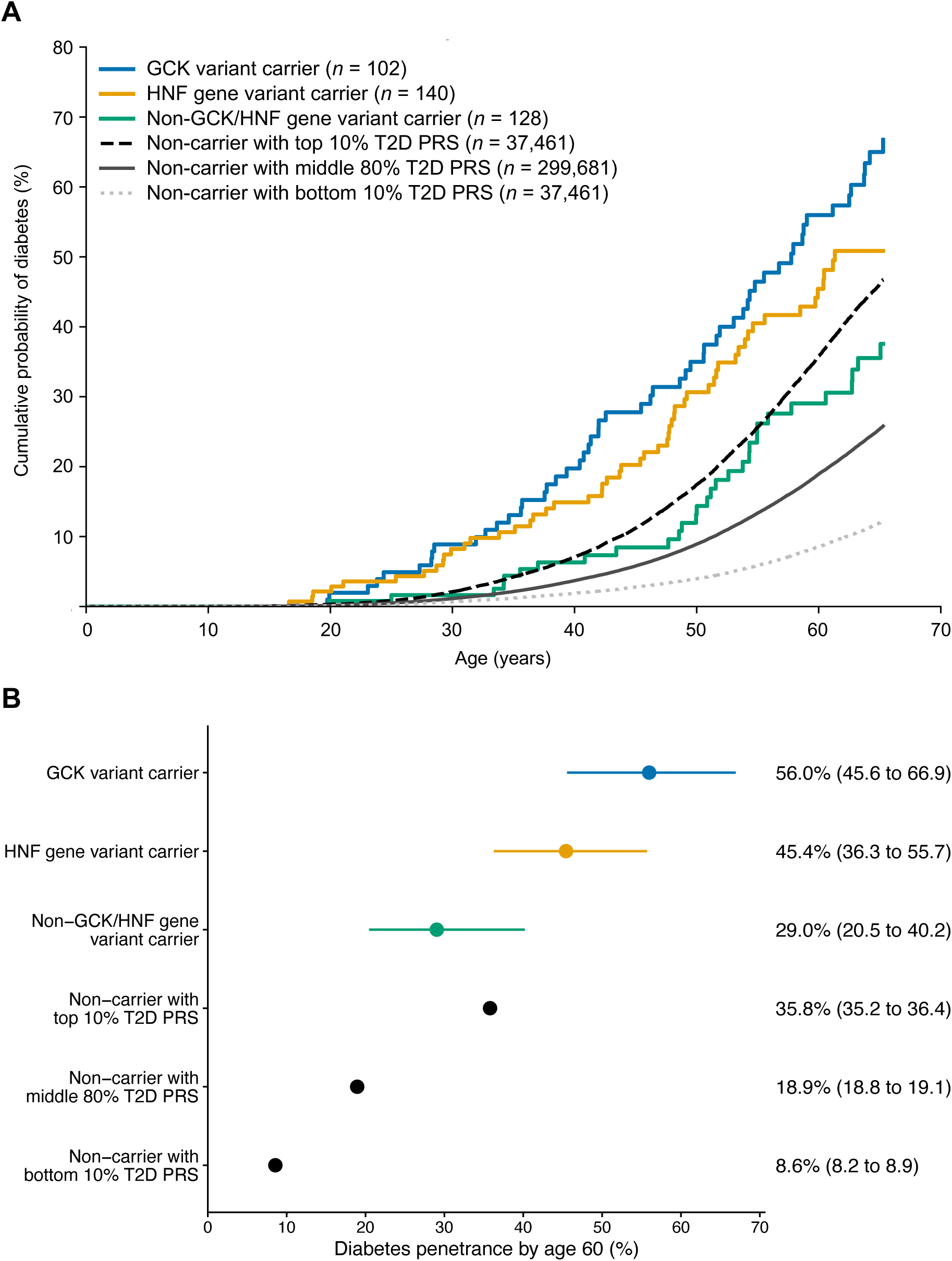
Diabetes penetrance among carriers of pathogenic or likely pathogenic variants in MODY genes and non-carriers stratified by type 2 diabetes polygenic risk score. Kaplan-Meier curves show the cumulative probability of diabetes by age for *GCK* variant carriers (*n* = 102), HNF gene variant carriers (*n* = 140), non-GCK/HNF gene variant carriers (*n* = 128), and non-carriers grouped by type 2 diabetes polygenic risk score (T2D PRS): top 10% (*n* = 37,461), middle 80% (*n* = 299,681), and bottom 10% (*n* = 37,461) (A). Using non-carriers in the middle 80% of the T2D PRS distribution as the reference, pairwise log-rank *P* values for the three carrier groups were 2.35 × 10^−33^, 1.39 × 10^−16^, and 0.014, respectively; *P* values for non-carriers in the top and bottom 10% of the T2D PRS distribution were both <1.0 × 10^−300^. Kaplan-Meier curves are truncated before the first age at which any displayed group has fewer than 20 participants at risk. Diabetes penetrance by age 60 is shown for each group (B). Points indicate Kaplan-Meier estimates, and horizontal bars indicate 95% confidence intervals (CIs). HNF genes include *HNF1A*, *HNF1B*, and *HNF4A*. Non-GCK/HNF genes include *ABCC8*, *INS*, *KCNJ11*, *NEUROD1*, *PDX1*, and *RFX6*.

Among participants with diabetes by age 40, 0.357% carried a P/LP variant in any evaluated MODY gene, corresponding to approximately 1 in 280 diabetes cases (**Supplementary Table 6**). Among participants with diabetes by age 60, the corresponding proportion was 0.253%, or approximately 1 in 395 cases. The medication-excluded definition yielded slightly higher carrier proportions, whereas the broader diabetes-label definition gave similar or slightly lower proportions than the primary definition (**Supplementary Table 6**).

### Age-Dependent Diabetes Onset by MODY Gene Group and T2D PRS

In Cox models using non-carriers in the middle group as the reference, HRs for diabetes onset were 4.06 (95% CI, 3.19 to 5.16) among *GCK* variant carriers, 2.72 (95% CI, 2.12 to 3.48) among HNF gene variant carriers, and 1.57 (95% CI, 1.14 to 2.17) among non-GCK/HNF gene variant carriers (**Figure 3A**). The HR for non-GCK/HNF gene variant carriers was close to that for non-carriers in the top 10% of T2D PRS (HR, 1.64; 95% CI, 1.60 to 1.68). Ancestry-stratified analyses showed broadly similar patterns in European and non-European ancestry groups, although 95% CIs for carrier groups were wider among non-European ancestry participants (**Figure 3B** and **3C**). Delayed-entry Cox models restricted to linked EHR follow-up yielded comparable findings (**Supplementary Figure 3**), and Schoenfeld residuals did not suggest clear violations of the proportional hazards assumption (**Supplementary Figure 4**).

**Figure 3.**
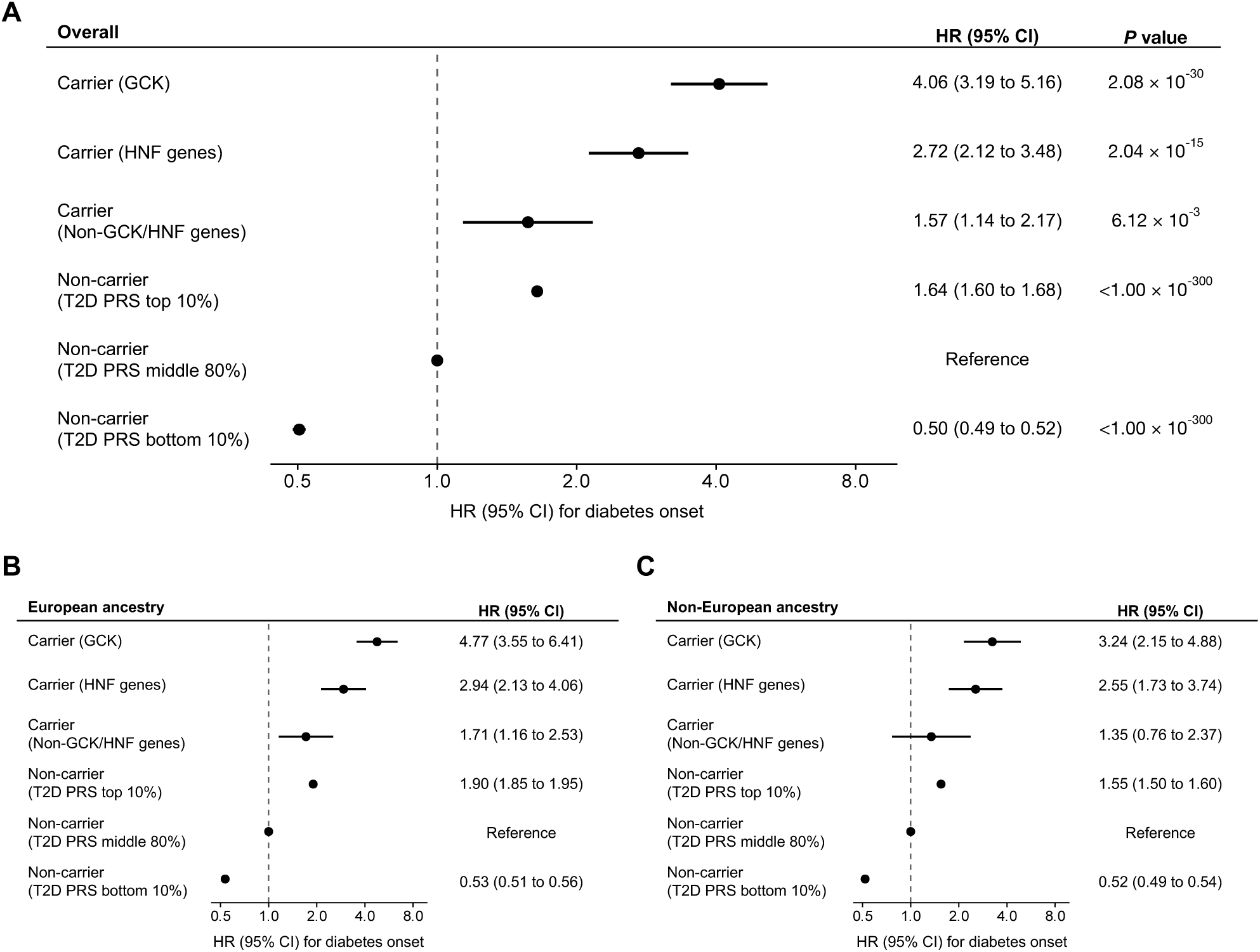
Diabetes onset in MODY gene variant carriers and non-carriers by type 2 diabetes polygenic risk score and genetic ancestry. Hazard ratios (HRs) for diabetes onset are shown overall (A), among European ancestry participants (B), and among non-European ancestry participants (C), from Cox proportional hazards models using age as the time scale. The overall analysis included carriers of pathogenic or likely pathogenic variants in *GCK* (*n* = 102), HNF genes (*n* = 140), and non-GCK/HNF genes (*n* = 128), and non-carriers in the top 10% (*n* = 37,461), middle 80% (*n* = 299,681; reference), and bottom 10% (*n* = 37,461) of the type 2 diabetes polygenic risk score (T2D PRS) distribution. Models were adjusted for sex and the first 10 genetic principal components. Points indicate HRs, and horizontal bars indicate 95% confidence intervals (CIs). HNF genes include *HNF1A*, *HNF1B*, and *HNF4A*. Non-GCK/HNF genes include *ABCC8*, *INS*, *KCNJ11*, *NEUROD1*, *PDX1*, and *RFX6*.

### Glycemic Profiles in MODY Gene Variant Carriers

Among participants with at least three HbA1c measurements, the analysis included 100 carriers and 71,280 non-carriers. Carriers included 37 with *GCK* variants, 41 with variants in HNF genes, and 22 with variants in non-GCK/HNF genes. Non-carriers included 8,712 in the top 10%, 56,822 in the middle group, and 5,746 in the bottom 10% of T2D PRS. *GCK* variant carriers showed relatively narrow HbA1c distributions, whereas HNF gene variant carriers, non-GCK/HNF gene variant carriers, and non-carriers in the top 10% of T2D PRS showed wider distributions, particularly for maximum HbA1c and HbA1c SD (**Figure 4**).

**Figure 4.**
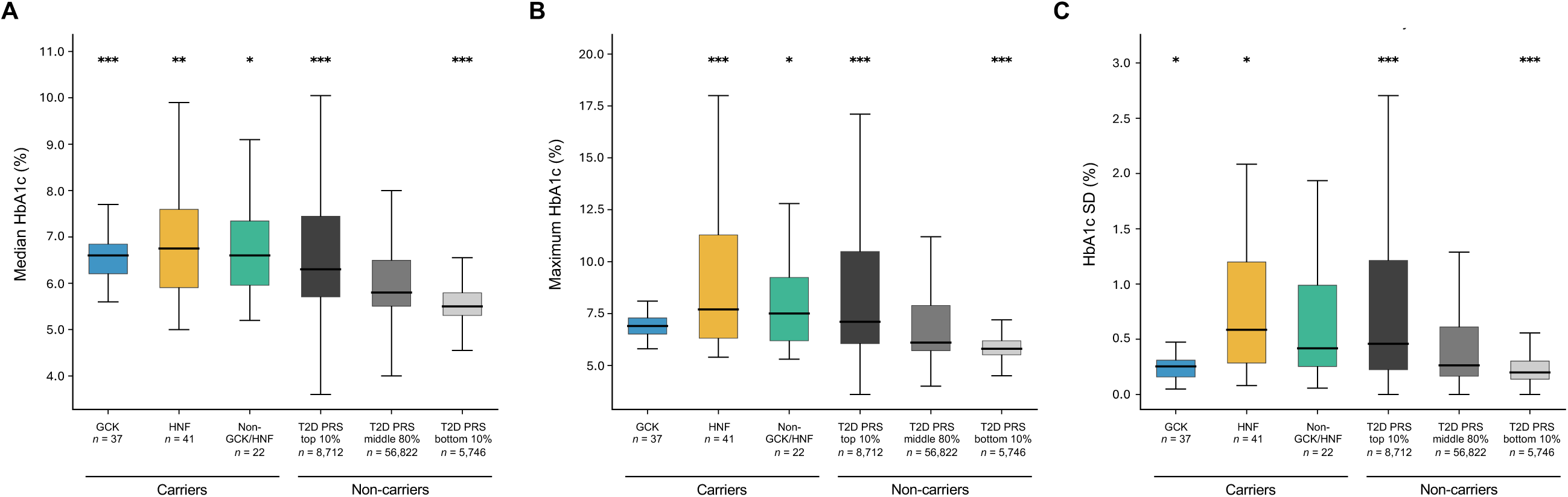
HbA1c profiles among MODY gene variant carriers and non-carriers stratified by type 2 diabetes polygenic risk score. Box plots show the distributions of median HbA1c (A), maximum HbA1c (B), and within-person HbA1c standard deviation (SD) (C) among participants with at least three HbA1c measurements separated by at least 90 days. Participants were grouped as *GCK* variant carriers (*n* = 37), HNF gene variant carriers (*n* = 41), non-GCK/HNF gene variant carriers (*n* = 22), and non-carriers in the top 10% (*n* = 8,712), middle 80% (*n* = 56,822), and bottom 10% (*n* = 5,746) of type 2 diabetes polygenic risk score (T2D PRS). Boxes indicate the interquartile range, with the horizontal line denoting the median. Whiskers extend to the most extreme values within 1.5 × the interquartile range; individual values beyond the whiskers are not displayed. \**P* < 0.05, \*\**P* < 0.01, and \*\*\**P* < 0.001 for comparisons with non-carriers in the middle 80% of T2D PRS, based on linear regression models adjusted for age at the midpoint of the HbA1c observation window, sex, and the first 10 genetic principal components. No symbol indicates *P* ≥ 0.05. HNF genes include *HNF1A*, *HNF1B*, and *HNF4A*. Non-GCK/HNF genes include *ABCC8*, *INS*, *KCNJ11*, *NEUROD1*, *PDX1*, and *RFX6*.

In linear regression models using non-carriers in the middle group as the reference, median HbA1c was higher in all carrier groups and in non-carriers in the top 10% of T2D PRS (**Figure 4A**). Adjusted differences were 0.55 percentage points for *GCK* variant carriers, 0.68 for HNF gene variant carriers, 0.74 for non-GCK/HNF gene variant carriers, and 0.53 for non-carriers in the top 10% of T2D PRS (all *P* < 0.05) (**Supplementary Table 7**). Maximum HbA1c was higher among HNF gene variant carriers, non-GCK/HNF gene variant carriers, and non-carriers in the top 10% of T2D PRS, with adjusted differences of 1.68, 1.08, and 0.97 percentage points, respectively, but not among *GCK* variant carriers (0.04 percentage points, *P* = 0.82) (**Figure 4B** and **Supplementary Table 7**). For HbA1c SD, adjusted differences were positive among HNF gene variant carriers and non-carriers in the top 10% of T2D PRS, negative among *GCK* variant carriers, and not clearly different among non-GCK/HNF gene variant carriers (**Figure 4C** and **Supplementary Table 7**).

## DISCUSSION

In this genotype-first study of 374,973 participants in the multi-ancestry *All of Us* Research Program (42.0% non-European ancestry), we identified 370 carriers of P/LP variants in 10 established MODY genes, corresponding to approximately 1 in 1,013 participants. Carriers were identified across ancestry groups, and MODY gene variants were associated with increased diabetes risk in both European and non-European ancestry groups. However, penetrance was incomplete, and age-dependent diabetes risk and glycemic profiles varied across and within genetic etiologies. When carriers were compared with non-carriers across the T2D PRS distribution, non-GCK/HNF gene variant carriers showed diabetes risk closer to that of non-carriers with high polygenic susceptibility than to that of *GCK* or HNF gene variant carriers. To our knowledge, this is the first study in a large, ancestrally diverse population to characterize P/LP MODY gene variant prevalence and carrier phenotypes using a genotype-first approach.

Carrier prevalence in this multi-ancestry study was nearly identical to that reported in the UK Biobank genotype-first study (1 in 1,013 vs. 1 in 1,052), and the overall gene distribution was comparable (10). Carrier prevalence was similar among European- and African-ancestry participants, the two largest ancestry groups (0.105% in both groups). The absence of female predominance among carriers was also consistent with a genotype-first approach, in contrast to phenotype-first studies, in which females have often been overrepresented (14, 17). Although overall prevalence was similar, the present study identified more distinct P/LP sequence variants than the UK Biobank study (202 vs. 182) despite the smaller sample size, a difference that may reflect the greater ancestral diversity of *All of Us* (10). Notably, 60 of 203 distinct P/LP variants (29.6%) were observed only among participants with non-European ancestry, highlighting the value of multi-ancestry cohorts for characterizing a broader variant spectrum of MODY genes. Although the test for heterogeneity suggested variation in carrier prevalence across ancestry groups, the ancestry-stratified findings should be interpreted cautiously because carrier counts were modest and variant-level evidence for pathogenicity classification has been derived disproportionately from European-ancestry populations (35).

Diabetes penetrance in this study was incomplete and differed by genetic etiology. At age 60, penetrance was 56.0%, 45.4%, and 29.0% among *GCK*, HNF, and non-GCK/HNF gene variant carriers, respectively. The lower value in the last group may have been influenced by *RFX6*, which represented nearly 70% of this category and has been associated with reduced-penetrance MODY (36). For *GCK*, the gap between diabetes penetrance in this study and the near-complete penetrance of mild hyperglycemia reported in previous studies (9) likely reflects EHR-based ascertainment, because mild hyperglycemia may not consistently be documented as diabetes in routine care. This ascertainment issue is also relevant to the observed proportion of MODY among diabetes cases in the current study, which was lower than in some population cohorts with standardized phenotyping (10). In EHR-linked cohorts, diabetes ascertainment relies on routine clinical documentation rather than standardized study measurements, which can capture a broad spectrum of clinically recognized diabetes while missing mild or asymptomatic hyperglycemia (37). A previous study showed a similar ascertainment pattern, with MODY representing a lower proportion of diabetes cases in an EHR-based cohort than in a population cohort with standardized phenotyping at baseline (38).

Comparing MODY gene variant carriers with non-carriers across the T2D PRS distribution helped interpret age-dependent diabetes risk and glycemic phenotypes in a population-wide context. Notably, non-carriers in the top 10% of T2D PRS, a much larger group than MODY gene variant carriers, had a hazard of diabetes onset comparable to that of non-GCK/HNF gene variant carriers. Similarly, among participants with repeated HbA1c measurements, non-carriers in the top 10% of T2D PRS showed broad HbA1c distributions and greater within-person variability, resembling the glycemic profiles of HNF and non-GCK/HNF gene variant carriers. These findings suggest that some MODY gene variant carrier phenotypes, particularly those associated with non-GCK/HNF genes, may overlap with phenotypes associated with high common-variant polygenic susceptibility. Prior studies have also shown that common-variant burden may contribute to a substantial fraction of early-onset diabetes with MODY-like features. For example, early-onset diabetes cases that clinically resembled MODY but had no identified monogenic cause had markedly higher T2D PRS (33). In a study of youth-onset type 2 diabetes, high diabetes risk driven primarily by common variants was more frequent than monogenic diabetes variants (12.6% vs. 2.4%) (39). At the population level, P/LP variants in MODY genes, particularly those in lower-penetrance genes, may confer increased but heterogeneous diabetes susceptibility that overlaps with high common-variant burden.

This study has several limitations. First, the number of carriers was limited, particularly when stratified by gene or ancestry. Some groups therefore had to be combined, limiting detailed assessment of gene- or ancestry-specific patterns. Second, the limited number of carriers also precluded evaluation of factors that may modify penetrance or glycemic heterogeneity among MODY gene variant carriers, including common polygenic background, environmental exposures, and treatment (40). Third, variant ascertainment and classification were intentionally conservative to minimize false-positive pathogenicity assignments, a major concern in population-scale genotype-first studies (5, 7). Some rare variants that were not reviewed or were not classified as P/LP after review may nevertheless contribute to diabetes susceptibility. Fourth, diabetes was ascertained from EHR-linked data, so age at ascertainment reflected the earliest available record rather than biological onset. Mild or asymptomatic hyperglycemia may have been missed or documented later than onset (37).

In summary, this genotype-first study of 374,973 *All of Us* participants identified P/LP variants in MODY genes across ancestry groups, with an overall prevalence of approximately 1 in 1,013 individuals and similar carrier prevalence among European- and African-ancestry participants. Although these variants were associated with increased diabetes risk, penetrance and glycemic profiles differed substantially by genetic etiology. Notably, diabetes risk among non-GCK/HNF gene variant carriers was comparable to that among non-carriers with high polygenic susceptibility, and glycemic profiles were broadly similar between these groups. Together, these results support equitable consideration of MODY genetic testing across ancestry groups and highlight the need for cautious clinical interpretation of P/LP variants in lower-penetrance genes beyond a single diagnostic label.

## Data Availability

Individual-level data are available to approved researchers through the All of Us Researcher Workbench under the All of Us Research Program data access policies. The authors are not permitted to redistribute participant-level data.

## Acknowledgments

We thank all participants in the *All of Us* Research Program, whose contributions made this study possible. We also thank the National Institutes of Health’s *All of Us* Research Program for providing access to the participant data analyzed in this study. M.H. is supported by the Japan Student Services Organization (JASSO; Graduate Scholarship for Degree-Seeking Study Abroad, selected under the special priority framework for doctoral studies at top-tier global universities in STEM fields) and the Watanabe Foundation (6th Toshizo Watanabe International Scholarship).

## Author Contributions

M.H. and S.Y. conceived and designed the study. M.H. processed, analyzed, and visualized the data and drafted the manuscript. M.H. and S.Y. manually reviewed and classified the variants included in the analysis and interpreted the results. S.Y. critically revised the manuscript for important intellectual content. Both authors reviewed and approved the final version of the manuscript. S.Y. is the guarantor of this work and, as such, had full access to all the data in the study and takes responsibility for the integrity of the data and the accuracy of the data analysis.

## Prior Presentation

None.

