## Supplementary Materials for "Population-Scale, Genotype-First Characterization of Monogenic Diabetes in 374,973 Multi-Ancestry Individuals from the *All of Us* Research Program"

### Contents


#### Supplementary Figures

Supplementary Figure 1. Diabetes penetrance among MODY gene variant carriers and non-carriers stratified by type 2 diabetes polygenic risk score.
Supplementary Figure 2. Kaplan-Meier curves for diabetes under alternative diabetes definitions.
Supplementary Figure 3. Delayed-entry Cox sensitivity analysis for diabetes onset in MODY variant carriers and non-carriers by type 2 diabetes polygenic risk score and genetic ancestry.
Supplementary Figure 4. Schoenfeld residual plots for Cox models of diabetes onset.

#### Supplementary Tables

Supplementary Table 1. Characteristics of *All of Us* participants by MODY gene variant carrier status.
Supplementary Table 2. Pathogenic and likely pathogenic variants in 10 established MODY genes identified in the *All of Us* Research Program.
Supplementary Table 3. Carrier prevalence and distinct pathogenic or likely pathogenic variant counts in MODY genes by sex and genetic ancestry.
Supplementary Table 4. Distribution of MODY gene variant carriers and distinct pathogenic or likely pathogenic variants by gene.
Supplementary Table 5. Kaplan-Meier-estimated diabetes penetrance under primary and alternative diabetes definitions.
Supplementary Table 6. Proportion of MODY gene variant carriers among participants with diabetes by ages 40 and 60.
Supplementary Table 7. HbA1c profiles among MODY gene variant carriers and non-carriers stratified by type 2 diabetes polygenic risk score.

#### Supplementary Notes

Supplementary Note 1. Variant classification.
Supplementary Note 2. Genotype and sequencing-read quality control for sequence variants.
Supplementary Note 3. Ascertainment of recurrent 17q12 deletions.
Supplementary Note 4. Diabetes ascertainment.
Supplementary Note References

**Supplementary Figure 1.** Diabetes penetrance among MODY gene variant carriers and non-carriers stratified by type 2 diabetes polygenic risk score.

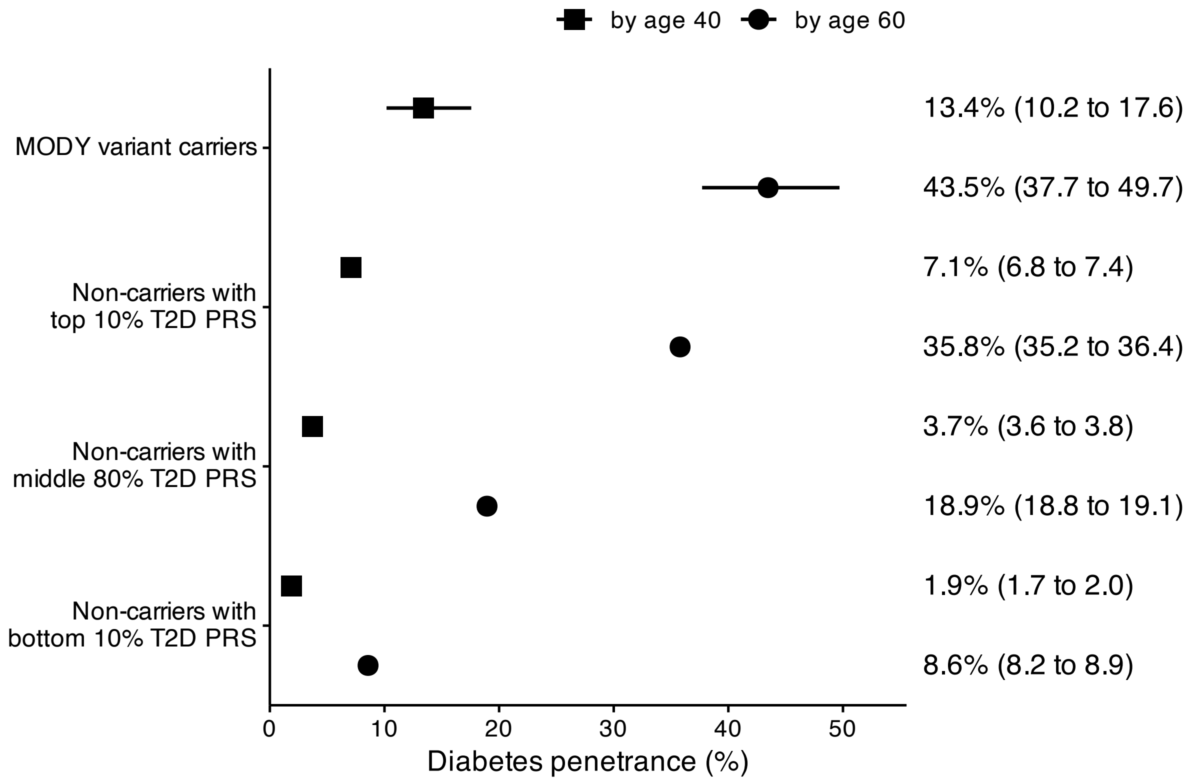


Kaplan-Meier diabetes penetrance by ages 40 and 60 is shown for all carriers of pathogenic or likely pathogenic variants in MODY genes (*n* = 370) and non-carriers stratified by type 2 diabetes polygenic risk score (T2D PRS): top 10% (*n* = 37,461), middle 80% (*n* = 299,681), and bottom 10% (*n* = 37,461), under the primary diabetes definition. Points indicate penetrance, and horizontal bars indicate 95% confidence intervals.

**Supplementary Figure 2.** Kaplan-Meier curves for diabetes under alternative diabetes definitions.

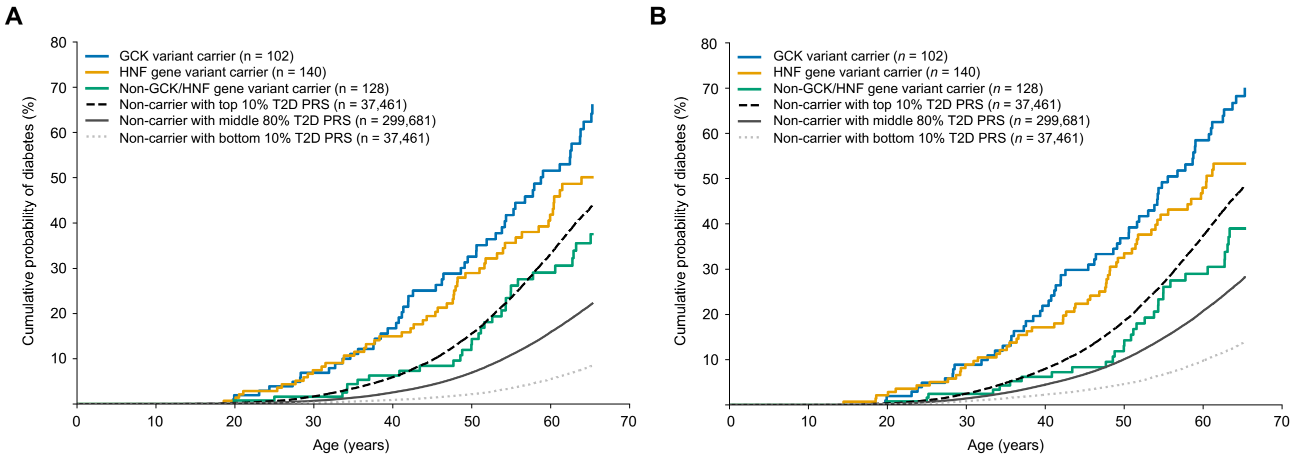


Kaplan-Meier curves show the cumulative probability of diabetes by age under the medication-excluded diabetes definition (A) and the broader diabetes-label definition (B). Curves are shown for *GCK* variant carriers (*n* = 102), HNF gene variant carriers (*n* = 140), non-GCK/HNF gene variant carriers (*n* = 128), and non-carriers grouped according to type 2 diabetes polygenic risk score (T2D PRS): top 10% (*n* = 37,461), middle 80% (*n* = 299,681), and bottom 10% (*n* = 37,461). Using non-carriers in the middle 80% of the T2D PRS distribution as the reference, pairwise log-rank *P* values for the three carrier groups were $5.22 \times 10^{-38}$, $8.99 \times 10^{-20}$, and $3.44 \times 10^{-4}$ in panel A, and $2.04 \times 10^{-32}$, $1.74 \times 10^{-16}$, and 0.058 in panel B, respectively; *P* values for non-carriers in the top and bottom 10% of the T2D PRS distribution were $<1.0 \times 10^{-300}$ in both panels. Kaplan-Meier curves are truncated before the first age at which any displayed group has fewer than 20 participants at risk. HNF genes include *HNF1A*, *HNF1B*, and *HNF4A*. Non-GCK/HNF genes include *ABCC8*, *INS*, *KCNJ11*, *NEUROD1*, *PDX1*, and *RFX6*.

**Supplementary Figure 3.** Delayed-entry Cox sensitivity analysis for diabetes onset in MODY variant carriers and non-carriers by type 2 diabetes polygenic risk score and genetic ancestry.

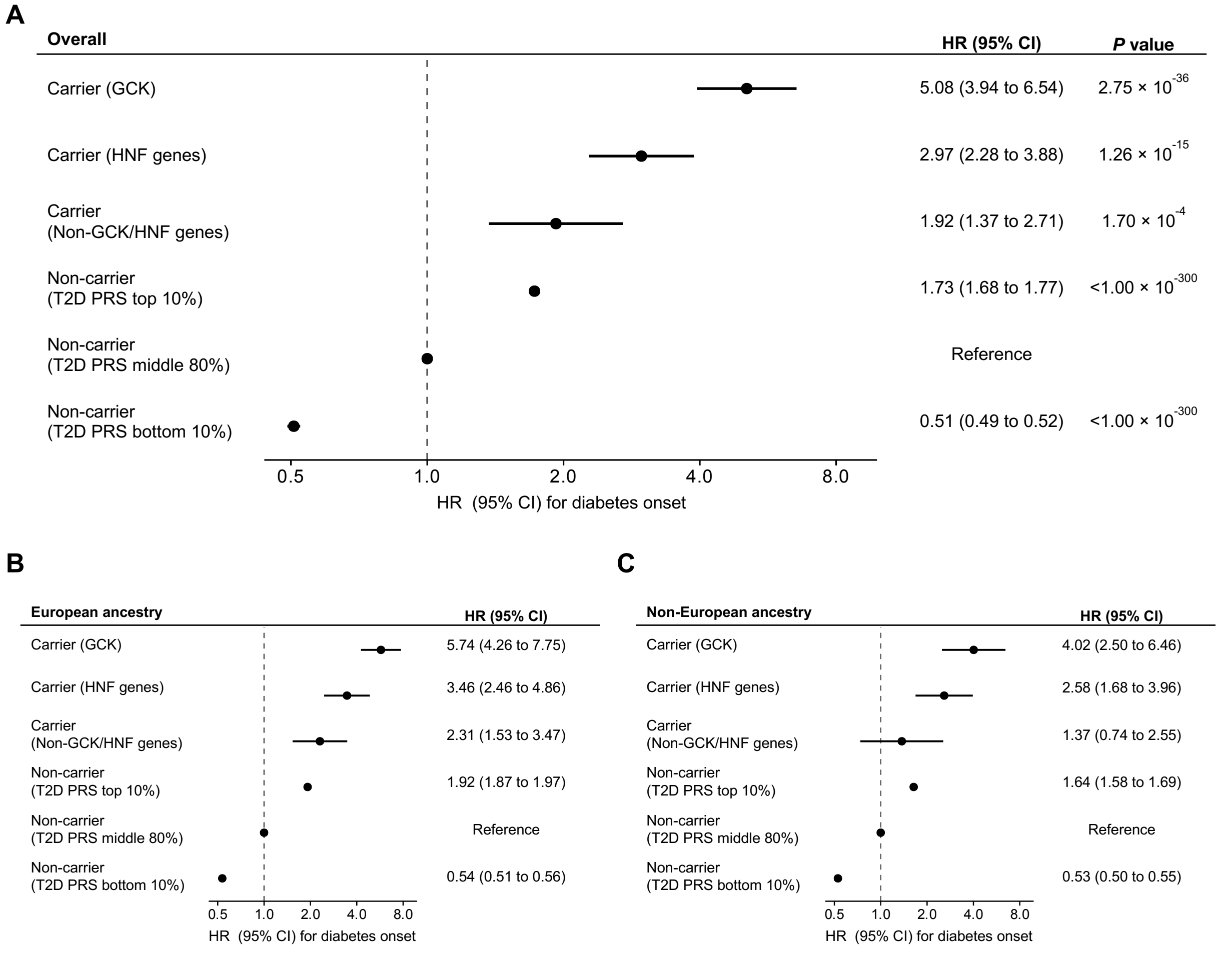


Hazard ratios (HRs) for diabetes onset are shown overall (A), among European ancestry participants (B), and among non-European ancestry participants (C), from delayed-entry Cox proportional hazards models using age as the time scale. The overall analysis included carriers of pathogenic or likely pathogenic variants in *GCK* (*n* = 89), HNF genes (*n* = 120), and non-GCK/HNF genes (*n* = 115), and non-carriers in the top 10% (*n* = 31,531), middle 80% (*n* = 272,402; reference), and bottom 10% (*n* = 35,590) of the type 2 diabetes polygenic risk score (T2D PRS) distribution. Entry was defined as age at the start of electronic health record observation, and exit as age at diabetes ascertainment during observation or at the end of observation. Participants with diabetes ascertained before the start of observation or without follow-up after observation began were excluded. Models were adjusted for sex and the first 10 genetic principal components. Points indicate HRs, and horizontal bars indicate 95% confidence intervals (CIs). HNF genes include *HNF1A*, *HNF1B*, and *HNF4A*. Non-GCK/HNF genes include *ABCC8*, *INS*, *KCNJ11*, *NEUROD1*, *PDX1*, and *RFX6*.

**Supplementary Figure 4**. Schoenfeld residual plots for Cox models of diabetes onset.

**A. Overall**

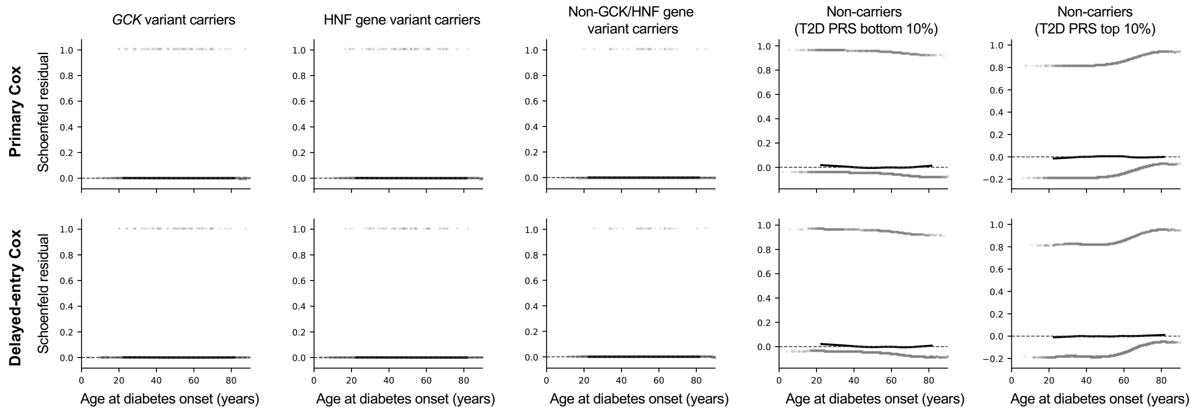


**B. European ancestry**

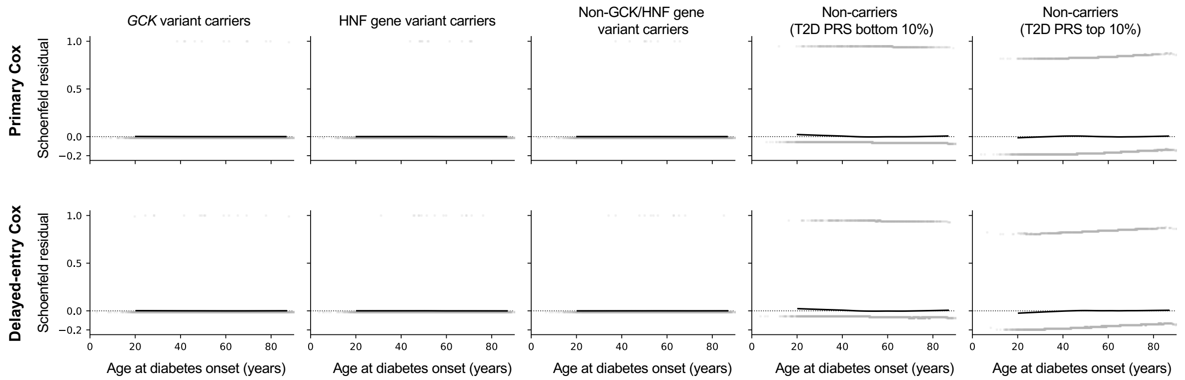


**C. Non-European ancestry**

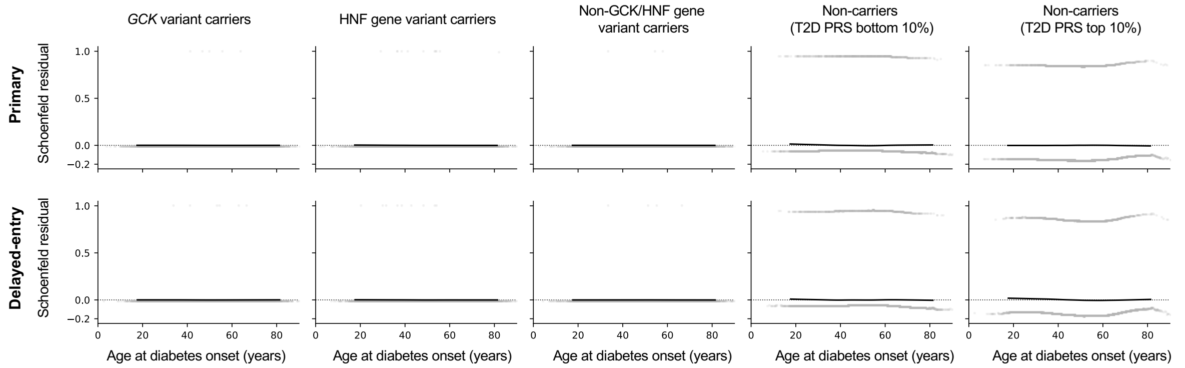


Scaled Schoenfeld residuals are shown for the overall population (A), European ancestry participants (B), and non-European ancestry participants (C), separately for the primary and delayed-entry Cox models. Models used age as the time scale and were adjusted for sex and the first 10 genetic principal components. The middle 80% of the type 2 diabetes polygenic risk score (T2D PRS) distribution among non-carriers was the reference group and is not shown. Gray points show scaled Schoenfeld residuals, black lines show smoothed residual trends, gray lines show uncertainty bands, and dotted horizontal lines indicate zero. HNF genes include *HNF1A*, *HNF1B*, and *HNF4A*. Non-GCK/HNF genes include *ABCC8*, *INS*, *KCNJ11*, *NEUROD1*, *PDX1*, and *RFX6*.

**Supplementary Table 1**. Characteristics of *All of Us* participants by MODY gene variant carrier status.

| Variable | Overall | Non-carrier | Carrier |
|---|---|---|---|
| Number of participants | 374,973 | 374,603 | 370 |
| Age at recruitment, years | 53.0 [37.0, 65.0] | 53.0 [37.0, 65.0] | 52.0 [37.0, 66.0] |
| Sex | | | |
| Female | 225,728 (60.2%) | 225,511 (60.2%) | 217 (58.6%) |
| Male | 149,245 (39.8%) | 149,092 (39.8%) | 153 (41.4%) |
| Ancestry | | | |
| African | 72,346 (19.3%) | 72,270 (19.3%) | 76 (20.5%) |
| Admixed American | 69,006 (18.4%) | 68,963 (18.4%) | 43 (11.6%) |
| European | 217,586 (58.0%) | 217,358 (58.0%) | 228 (61.6%) |
| Other ancestry | 16,035 (4.3%) | 16,012 (4.3%) | 23 (6.2%) |

Data are presented as median [IQR] for age at recruitment and *n* (%) for categorical variables. Percentages were calculated within each column. Carrier status was defined by the presence of a pathogenic or likely pathogenic variant in any of the 10 MODY genes evaluated. Genetic ancestry was based on *All of Us* genetic ancestry assignments and is reported as genetic ancestry groups. In the non-carrier and carrier columns, South Asian, East Asian, and Middle Eastern groups were combined to avoid reporting cells with fewer than 20 participants.

**Supplementary Table 2**. Pathogenic and likely pathogenic variants in 10 established MODY genes identified in the *All of Us* Research Program.

| Gene | Transcript | Canonical SPDI / variant representation | cDNA | Protein | Effect | Pathogenicity |
|---|---|---|---|---|---|---|
| *ABCC8* | NM_000352.6 | NC_000011.10:17394267:G:A | c.4544C>T | p.Thr1515Met | missense | Pathogenic |
| *ABCC8* | NM_000352.6 | NC_000011.10:17397788:C:T | c.3763G>A | p.Gly1255Ser | missense | Likely pathogenic |
| *ABCC8* | NM_000352.6 | NC_000011.10:17404523:C:T | c.3545G>A | p.Arg1182Gln | missense | Pathogenic |
| *ABCC8* | NM_000352.6 | NC_000011.10:17413395:G:A | c.2473C>T | p.Arg825Trp | missense | Pathogenic |
| *ABCC8* | NM_000352.6 | NC_000011.10:17460581:C:T | c.917G>A | p.Arg306His | missense | Pathogenic |
| *ABCC8* | NM_000352.6 | NC_000011.10:17461782:C:T | c.622G>A | p.Glu208Lys | missense | Likely pathogenic |
| *GCK* | NM_000162.5 | NC_000007.14:44145280:C:A | c.1254-1G>T | – | splice acceptor | Pathogenic |
| *GCK* | NM_000162.5 | NC_000007.14:44145494:A:G | c.1253+2T>C | – | splice donor | Pathogenic |
| *GCK* | NM_000162.5 | NC_000007.14:44145562:CTCTC:CTCTCTC | c.1186_1187dup | p.Ser396fs | frameshift | Pathogenic |
| *GCK* | NM_000162.5 | NC_000007.14:44145566:C:A | c.1183G>T | p.Glu395Ter | stop gained | Pathogenic |
| *GCK* | NM_000162.5 | NC_000007.14:44145572:T: | c.1177del | p.Met393Cysfs | frameshift | Pathogenic |
| *GCK* | NM_000162.5 | NC_000007.14:44145594:CCCC:CCC | c.1155del | p.Leu386fs | frameshift | Pathogenic |
| *GCK* | NM_000162.5 | NC_000007.14:44145670:G:T | c.1079C>A | p.Ser360Ter | stop gained | Pathogenic |
| *GCK* | NM_000162.5 | NC_000007.14:44145695:GG:G | c.1054del | p.Ile351_Leu352insTer | stop gained | Pathogenic |
| *GCK* | NM_000162.5 | NC_000007.14:44146618:C:T | c.864-1G>A | – | splice acceptor | Pathogenic |
| *GCK* | NM_000162.5 | NC_000007.14:44147834:T:C | c.680-2A>G | – | splice acceptor | Pathogenic |
| *GCK* | NM_000162.5 | NC_000007.14:44149772:G: | c.666del | p.Gly223fs | frameshift | Pathogenic |
| *GCK* | NM_000162.5 | NC_000007.14:44149778:G:T | c.660C>A | p.Cys220Ter | stop gained | Pathogenic |

| | | | | | | |
|---|---|---|---|---|---|---|
| *GCK* | NM_000162.5 | NC_000007.14:44149793:G:C | c.645C>G | p.Tyr215Ter | stop gained | Pathogenic |
| *GCK* | NM_000162.5 | NC_000007.14:44149793:G:T | c.645C>A | p.Tyr215Ter | stop gained | Pathogenic |
| *GCK* | NM_000162.5 | NC_000007.14:44149859:C:G | c.580-1G>C | – | splice acceptor | Pathogenic |
| *GCK* | NM_000162.5 | NC_000007.14:44149980:GATAG: GATAGATAG | c.564_567dup | p.Lys190fs | frameshift | Pathogenic |
| *GCK* | NM_000162.5 | NC_000007.14:44149991:G:A | c.556C>T | p.Arg186Ter | stop gained | Pathogenic |
| *GCK* | NM_000162.5 | NC_000007.14:44150065:T:C | c.484-2A>G | – | splice acceptor | Pathogenic |
| *GCK* | NM_000162.5 | NC_000007.14:44150977:A: | c.461delT | p.Val154Glyfs | frameshift | Pathogenic |
| *GCK* | NM_000162.5 | NC_000007.14:44152425:C:T | c.209-1G>A | – | splice acceptor | Pathogenic |
| *GCK* | NM_000162.5 | NC_000007.14:44152426:T:C | c.209-2A>G | – | splice acceptor | Pathogenic |
| *GCK* | NM_000162.5 | NC_000007.14:44153325:G:T | c.183C>A | p.Tyr61Ter | stop gained | Pathogenic |
| *GCK* | NM_000162.5 | NC_000007.14:44188907:C:T | c.45+1G>A | – | splice donor | Pathogenic |
| *GCK* | NM_000162.5 | NC_000007.14:44145175:G:A | c.1358C>T | p.Ser453Leu | missense | Pathogenic |
| *GCK* | NM_000162.5 | NC_000007.14:44145193:C:T | c.1340G>A | p.Arg447Gln | missense | Likely pathogenic |
| *GCK* | NM_000162.5 | NC_000007.14:44145269:G:A | c.1264C>T | p.Arg422Trp | missense | Likely pathogenic |
| *GCK* | NM_000162.5 | NC_000007.14:44145515:C:T | c.1234G>A | p.Val412Met | missense | Pathogenic |
| *GCK* | NM_000162.5 | NC_000007.14:44145521:C:G | c.1228G>C | p.Gly410Arg | missense | Pathogenic |
| *GCK* | NM_000162.5 | NC_000007.14:44145596:C:T | c.1153G>A | p.Gly385Arg | missense | Likely pathogenic |
| *GCK* | NM_000162.5 | NC_000007.14:44145613:G:T | c.1136C>A | p.Ala379Glu | missense | Likely pathogenic |
| *GCK* | NM_000162.5 | NC_000007.14:44145617:C:T | c.1132G>A | p.Ala378Thr | missense | Pathogenic |
| *GCK* | NM_000162.5 | NC_000007.14:44145619:C:T | c.1130G>A | p.Arg377His | missense | Likely pathogenic |
| *GCK* | NM_000162.5 | NC_000007.14:44145662:C:T | c.1087G>A | p.Asp363Asn | missense | Likely pathogenic |
| *GCK* | NM_000162.5 | NC_000007.14:44146462:C:T | c.1019G>A | p.Ser340Asn | missense | Pathogenic |

| | | | | | | |
|---|---|---|---|---|---|---|
| *GCK* | NM_000162.5 | NC_000007.14:44146534:A:T | c.947T>A | p.Phe316Tyr | missense | Likely pathogenic |
| *GCK* | NM_000162.5 | NC_000007.14:44147678:G:C | c.834C>G | p.Asp278Glu | missense | Pathogenic |
| *GCK* | NM_000162.5 | NC_000007.14:44147691:T:G | c.821A>C | p.Asp274Ala | missense | Pathogenic |
| *GCK* | NM_000162.5 | NC_000007.14:44147722:C:T | c.790G>A | p.Gly264Ser | missense | Pathogenic |
| *GCK* | NM_000162.5 | NC_000007.14:44147748:G:A | c.764C>T | p.Thr255Ile | missense | Pathogenic |
| *GCK* | NM_000162.5 | NC_000007.14:44147754:A:G | c.758T>C | p.Val253Ala | missense | Likely pathogenic |
| *GCK* | NM_000162.5 | NC_000007.14:44147760:A:G | c.752T>C | p.Met251Thr | missense | Likely pathogenic |
| *GCK* | NM_000162.5 | NC_000007.14:44147763:C:T | c.749G>A | p.Arg250His | missense | Likely pathogenic |
| *GCK* | NM_000162.5 | NC_000007.14:44147764:G:A | c.748C>T | p.Arg250Cys | missense | Pathogenic |
| *GCK* | NM_000162.5 | NC_000007.14:44147775:C:T | c.737G>A | p.Gly246Glu | missense | Likely pathogenic |
| *GCK* | NM_000162.5 | NC_000007.14:44147808:A:G | c.704T>C | p.Met235Thr | missense | Pathogenic |
| *GCK* | NM_000162.5 | NC_000007.14:44149821:G:A | c.617C>T | p.Thr206Met | missense | Pathogenic |
| *GCK* | NM_000162.5 | NC_000007.14:44149822:T:G | c.616A>C | p.Thr206Pro | missense | Pathogenic |
| *GCK* | NM_000162.5 | NC_000007.14:44149830:A:G | c.608T>C | p.Val203Ala | missense | Pathogenic |
| *GCK* | NM_000162.5 | NC_000007.14:44149833:A:G | c.605T>C | p.Met202Thr | missense | Pathogenic |
| *GCK* | NM_000162.5 | NC_000007.14:44149837:C:A | c.601G>T | p.Ala201Ser | missense | Pathogenic |
| *GCK* | NM_000162.5 | NC_000007.14:44149975:C:T | c.572G>A | p.Arg191Gln | missense | Pathogenic |
| *GCK* | NM_000162.5 | NC_000007.14:44149976:G:A | c.571C>T | p.Arg191Trp | missense | Pathogenic |
| *GCK* | NM_000162.5 | NC_000007.14:44150003:C:T | c.544G>A | p.Val182Met | missense | Pathogenic |
| *GCK* | NM_000162.5 | NC_000007.14:44150007:A:C | c.540T>G | p.Asn180Lys | missense | Likely pathogenic |
| *GCK* | NM_000162.5 | NC_000007.14:44150056:A:G | c.491T>C | p.Leu164Pro | missense | Pathogenic |
| *GCK* | NM_000162.5 | NC_000007.14:44150960:C:T | c.478G>A | p.Asp160Asn | missense | Pathogenic |

| | | | | | | |
|---|---|---|---|---|---|---|
| *GCK* | NM_000162.5 | NC_000007.14:44150969:C:T | c.469G>A | p.Glu157Lys | missense | Pathogenic |
| *GCK* | NM_000162.5 | NC_000007.14:44151028:T:C | c.410A>G | p.His137Arg | missense | Likely pathogenic |
| *GCK* | NM_000162.5 | NC_000007.14:44151068:C:T | c.370G>A | p.Asp124Asn | missense | Pathogenic |
| *GCK* | NM_000162.5 | NC_000007.14:44151074:G:T | c.364C>A | p.Leu122Ile | missense | Likely pathogenic |
| *GCK* | NM_000162.5 | NC_000007.14:44152394:C:G | c.239G>C | p.Gly80Ala | missense | Likely pathogenic |
| *GCK* | NM_000162.5 | NC_000007.14:44152419:C:T | c.214G>A | p.Gly72Arg | missense | Pathogenic |
| *GCK* | NM_000162.5 | NC_000007.14:44153323:A:G | c.185T>C | p.Val62Ala | missense | Pathogenic |
| *GCK* | NM_000162.5 | NC_000007.14:44153337:C:T | c.171G>A | p.Met57Ile | missense | Pathogenic |
| *GCK* | NM_000162.5 | NC_000007.14:44153350:G:A | c.158C>T | p.Ala53Val | missense | Likely pathogenic |
| *GCK* | NM_000162.5 | NC_000007.14:44153362:G:T | c.146C>A | p.Thr49Asn | missense | Likely pathogenic |
| *GCK* | NM_000162.5 | NC_000007.14:44153380:C:T | c.128G>A | p.Arg43His | missense | Pathogenic |
| *GCK* | NM_000162.5 | NC_000007.14:44153386:A:G | c.122T>C | p.Met41Thr | missense | Likely pathogenic |
| *HNF1A* | NM_000545.8 | NC_000012.12:120979029:G:T | c.262G>T | p.Glu88Ter | stop gained | Pathogenic |
| *HNF1A* | NM_000545.8 | NC_000012.12:120979058:G: | c.292del | p.Ala98fs | frameshift | Pathogenic |
| *HNF1A* | NM_000545.8 | NC_000012.12:120979094:G:A | c.326+1G>A | – | splice donor | Pathogenic |
| *HNF1A* | NM_000545.8 | NC_000012.12:120988830:A:C | c.327-2A>C | – | splice acceptor | Pathogenic |
| *HNF1A* | NM_000545.8 | NC_000012.12:120988875:C:T | c.370C>T | p.Gln124Ter | stop gained | Pathogenic |
| *HNF1A* | NM_000545.8 | NC_000012.12:120989013:C:T | c.508C>T | p.Gln170Ter | stop gained | Pathogenic |
| *HNF1A* | NM_000545.8 | NC_000012.12:120989031:C:T | c.526C>T | p.Gln176Ter | stop gained | Pathogenic |
| *HNF1A* | NM_000545.8 | NC_000012.12:120989032:G:A | c.526+1G>A | – | splice donor | Pathogenic |
| *HNF1A* | NM_000545.8 | NC_000012.12:120993677:C:T | c.685C>T | p.Arg229Ter | stop gained | Pathogenic |
| *HNF1A* | NM_000545.8 | NC_000012.12:120994204:AG: | c.755_756del | p.Gln252fs | frameshift | Pathogenic |

| | | | | | | |
|---|---|---|---|---|---|---|
| *HNF1A* | NM_000545.8 | NC_000012.12:120994311:GGG:GG | c.864del | p.Pro291fs | frameshift | Pathogenic |
| *HNF1A* | NM_000545.8 | NC_000012.12:120994313::C | c.863_864insC | p.Pro289fs | frameshift | Pathogenic |
| *HNF1A* | NM_000545.8 | NC_000012.12:120994314:CCCCCCCC:CCCCCCCCC | c.872dup | p.Gly292fs | frameshift | Pathogenic |
| *HNF1A* | NM_000545.8 | NC_000012.12:120994314:CCCCCCCC:CCCCCCC | c.872del | p.Pro291fs | frameshift | Pathogenic |
| *HNF1A* | NM_000545.8 | NC_000012.12:120994406:T:TT | c.955+2dup | – | splice donor | Pathogenic |
| *HNF1A* | NM_000545.8 | NC_000012.12:120994405:G:T | c.955+1G>T | – | splice donor | Pathogenic |
| *HNF1A* | NM_000545.8 | NC_000012.12:120996302:GTACCCTCAAGCAGCGG:G | c.998_1013del | p.Val333fs | frameshift | Pathogenic |
| *HNF1A* | NM_000545.8 | NC_000012.12:120996568:CT: | c.1136_1137del | p.Pro379fs | frameshift | Pathogenic |
| *HNF1A* | NM_000545.8 | NC_000012.12:120996687:CCTGCCTC: | c.1259_1266delCCTCCCTG | p.Ala420fs | frameshift | Pathogenic |
| *HNF1A* | NM_000545.8 | NC_000012.12:120997491:CACA:CA | c.1330_1331del | p.Gln444fs | frameshift | Pathogenic |
| *HNF1A* | NM_000545.8 | NC_000012.12:120999626:G: | c.1768del | p.Val590fs | frameshift | Pathogenic |
| *HNF1A* | NM_000545.8 | NC_000012.12:121001097:C: | c.1802del | p.Ser600_Ser601insTer | stop gained | Pathogenic |
| *HNF1A* | NM_000545.8 | NC_000012.12:121001114:C:T | c.1819C>T | p.Gln607Ter | stop gained | Pathogenic |
| *HNF1A* | NM_000545.8 | NC_000012.12:120978801:C:T | c.34C>T | p.Leu12Phe | missense | Likely pathogenic |
| *HNF1A* | NM_000545.8 | NC_000012.12:120978865:C:T | c.98C>T | p.Pro33Leu | missense | Likely pathogenic |
| *HNF1A* | NM_000545.8 | NC_000012.12:120988852:C:T | c.347C>T | p.Ala116Val | missense | Pathogenic |
| *HNF1A* | NM_000545.8 | NC_000012.12:120988896:C:T | c.391C>T | p.Arg131Trp | missense | Pathogenic |
| *HNF1A* | NM_000545.8 | NC_000012.12:120988897:G:A | c.392G>A | p.Arg131Gln | missense | Pathogenic |
| *HNF1A* | NM_000545.8 | NC_000012.12:120988980:C:T | c.475C>T | p.Arg159Trp | missense | Pathogenic |
| *HNF1A* | NM_000545.8 | NC_000012.12:120993590:C:T | c.598C>T | p.Arg200Trp | missense | Pathogenic |
| *HNF1A* | NM_000545.8 | NC_000012.12:120993600:G:A | c.608G>A | p.Arg203His | missense | Pathogenic |

| | | | | | | |
|---|---|---|---|---|---|---|
| *HNF1A* | NM_000545.8 | NC_000012.12:120993618:C:A | c.626C>A | p.Ala209Glu | missense | Likely pathogenic |
| *HNF1A* | NM_000545.8 | NC_000012.12:120993678:G:A | c.686G>A | p.Arg229Gln | missense | Likely pathogenic |
| *HNF1A* | NM_000545.8 | NC_000012.12:120994236:C:T | c.787C>T | p.Arg263Cys | missense | Pathogenic |
| *HNF1A* | NM_000545.8 | NC_000012.12:120994263:C:T | c.814C>T | p.Arg272Cys | missense | Pathogenic |
| *HNF1A* | NM_000545.8 | NC_000012.12:120994276:C:G | c.827C>G | p.Ala276Gly | missense | Likely pathogenic |
| *HNF1A* | NM_000545.8 | NC_000012.12:120994282:G:A | c.833G>A | p.Arg278Gln | missense | Likely pathogenic |
| *HNF1A* | NM_000545.8 | NC_000012.12:120997503:C:T | c.1340C>T | p.Pro447Leu | missense | Pathogenic |
| *HNF1B* | NM_000458.4 | 17q12 recurrent deletion involving *HNF1B* | whole-gene deletion | – | whole-gene deletion | Pathogenic |
| *HNF1B* | NM_000458.4 | NC_000017.11:37699131:CCAT: | c.1594_1597del | p.Met532Trpfs | frameshift | Pathogenic |
| *HNF1B* | NM_000458.4 | NC_000017.11:37699167:GGGGGGG:GGGGGGGG | c.1561dup | p.Gln521fs | frameshift | Pathogenic |
| *HNF1B* | NM_000458.4 | NC_000017.11:37699167:GGGGGGG:GGGGGG | c.1561del | p.Gln521fs | frameshift | Pathogenic |
| *HNF1B* | NM_000458.4 | NC_000017.11:37699179:G:A | c.1549C>T | p.Gln517Ter | stop gained | Pathogenic |
| *HNF1B* | NM_000458.4 | NC_000017.11:37731633:GGGGGG:GGGGGGG | c.1006dup | p.His336fs | frameshift | Pathogenic |
| *HNF1B* | NM_000458.4 | NC_000017.11:37733555:C:G | c.809+1G>C | – | splice donor | Pathogenic |
| *HNF1B* | NM_000458.4 | NC_000017.11:37733623:G:A | c.742C>T | p.Gln248Ter | stop gained | Pathogenic |
| *HNF1B* | NM_000458.4 | NC_000017.11:37733713:G:A | c.652C>T | p.Gln218Ter | stop gained | Pathogenic |
| *HNF1B* | NM_000458.4 | chr17:37731774-37731782 (collapsed local complex indel) | NA | – | frameshift | Pathogenic |
| *HNF1B* | NM_000458.4 | NC_000017.11:37710653:T:C | c.1055A>G | p.Tyr352Cys | missense | Likely pathogenic |
| *HNF1B* | NM_000458.4 | NC_000017.11:37733661:C:T | c.704G>A | p.Arg235Gln | missense | Likely pathogenic |
| *HNF1B* | NM_000458.4 | NC_000017.11:37733667:C:T | c.698G>A | p.Arg233His | missense | Likely pathogenic |
| *HNF1B* | NM_000458.4 | NC_000017.11:37739466:C:G | c.517G>C | p.Val173Leu | missense | Likely pathogenic |

| | | | | | | |
|---|---|---|---|---|---|---|
| *HNF4A* | NM_000457.6 | NC_000020.11:44414593:C:T | c.580C>T | p.Gln194Ter | stop gained | Pathogenic |
| *HNF4A* | NM_000457.6 | NC_000020.11:44418507:C: | c.733del | p.Leu245fs | frameshift | Pathogenic |
| *HNF4A* | NM_000457.6 | NC_000020.11:44424255::T | c.1129+1_1129+2insT | – | splice donor | Pathogenic |
| *HNF4A* | NM_000457.6 | NC_000020.11:44428332:A:G | c.1130-1A>G | – | splice acceptor | Pathogenic |
| *HNF4A* | NM_000457.6 | NC_000020.11:44428356:T:TT | c.1152dup | p.Ala385fs | frameshift | Pathogenic |
| *HNF4A* | NM_000457.6 | NC_000020.11:44428401:A: | c.1198del | p.Thr400Profs | frameshift | Pathogenic |
| *HNF4A* | NM_000457.6 | NC_000020.11:44428450:C:T | c.1246C>T | p.Gln416Ter | stop gained | Pathogenic |
| *HNF4A* | NM_000457.6 | NC_000020.11:44428468:C:T | c.1264C>T | p.Arg422Ter | stop gained | Likely pathogenic |
| *HNF4A* | NM_000457.6 | NC_000020.11:44429521:G:T | c.1283-1G>T | – | splice acceptor | Pathogenic |
| *HNF4A* | NM_000457.6 | NC_000020.11:44406131:G:A | c.190G>A | p.Gly64Arg | missense | Likely pathogenic |
| *HNF4A* | NM_000457.6 | NC_000020.11:44407410:C:A | c.321C>A | p.Asp107Glu | missense | Likely pathogenic |
| *HNF4A* | NM_000457.6 | NC_000020.11:44413707:C:T | c.400C>T | p.Arg134Trp | missense | Pathogenic |
| *HNF4A* | NM_000457.6 | NC_000020.11:44413708:G:A | c.401G>A | p.Arg134Gln | missense | Pathogenic |
| *HNF4A* | NM_000457.6 | NC_000020.11:44418455:A:C | c.680A>C | p.His227Pro | missense | Likely pathogenic |
| *HNF4A* | NM_000457.6 | NC_000020.11:44419741:G:A | c.758G>A | p.Arg253Gln | missense | Pathogenic |
| *HNF4A* | NM_000457.6 | NC_000020.11:44424121:C:T | c.997C>T | p.Arg333Cys | missense | Pathogenic |
| *HNF4A* | NM_000457.6 | NC_000020.11:44424122:G:A | c.998G>A | p.Arg333His | missense | Pathogenic |
| *INS* | NM_000207.3 | NC_000011.10:2160824:G:C | c.147C>G | p.Phe49Leu | missense | Pathogenic |
| *INS* | NM_000207.3 | NC_000011.10:2160877:C:T | c.94G>A | p.Gly32Ser | missense | Pathogenic |
| *KCNJ11* | NM_000525.4 | NC_000011.10:17387490:G:A | c.601C>T | p.Arg201Cys | missense | Pathogenic |
| *NEUROD1* | NM_002500.5 | NC_000002.12:181678199:A: | c.661del | p.Tyr221Thrfs | frameshift | Pathogenic |
| *NEUROD1* | NM_002500.5 | NC_000002.12:181678244:GGGGGGG:GGGGGGGG | c.616dup | p.His206fs | frameshift | Pathogenic |

| | | | | | | |
|---|---|---|---|---|---|---|
| *NEUROD1* | NM_002500.5 | NC_000002.12:181678535:GGG:GGGG | c.325dup | p.Arg109Profs | frameshift | Pathogenic |
| *NEUROD1* | NM_002500.5 | NC_000002.12:181678795:C:T | c.65G>A | p.Trp22Ter | stop gained | Pathogenic |
| *PDX1* | NM_000209.4 | NC_000013.11:27920320:CCCCCC:CCCCC | c.188del | p.Pro63fs | frameshift | Pathogenic |
| *PDX1* | NM_000209.4 | NC_000013.11:27924380:G:A | c.532G>A | p.Glu178Lys | missense | Likely pathogenic |
| *RFX6* | NM_173560.4 | NC_000006.12:116877303:CCTTCCTGCAGGCGCAG: | c.32_48del | p.Phe11CysfsTer | frameshift | Pathogenic |
| *RFX6* | NM_173560.4 | NC_000006.12:116877347:C:T | c.73C>T | p.Gln25Ter | stop gained | Pathogenic |
| *RFX6* | NM_173560.4 | NC_000006.12:116877434:G: | c.164del | p.Gly55Alafs | frameshift | Pathogenic |
| *RFX6* | NM_173560.4 | NC_000006.12:116877465:A: | c.192del | p.Asp65Thrfs | frameshift | Pathogenic |
| *RFX6* | NM_173560.4 | NC_000006.12:116877474::AGCTGCC | c.200_206dup | p.Gly70Alafs | frameshift | Pathogenic |
| *RFX6* | NM_173560.4 | NC_000006.12:116877481::G | c.211dup | p.Ala71Glyfs | frameshift | Pathogenic |
| *RFX6* | NM_173560.4 | NC_000006.12:116877815:AA:A | c.245del | p.Asn82fs | frameshift | Pathogenic |
| *RFX6* | NM_173560.4 | NC_000006.12:116877852:AC: | c.282_283del | p.Asp94Glufs | frameshift | Pathogenic |
| *RFX6* | NM_173560.4 | NC_000006.12:116877919::T | c.348dup | p.Lys117Ter | frameshift | Pathogenic |
| *RFX6* | NM_173560.4 | NC_000006.12:116877932:AC: | c.364_365del | p.Gln122Alafs | frameshift | Pathogenic |
| *RFX6* | NM_173560.4 | NC_000006.12:116880621:T:A | c.459T>A | p.Cys153Ter | stop gained | Pathogenic |
| *RFX6* | NM_173560.4 | NC_000006.12:116894038:G: | c.619del | p.Val207fs | frameshift | Pathogenic |
| *RFX6* | NM_173560.4 | NC_000006.12:116895177:A:G | c.645-2A>G | – | splice acceptor | Pathogenic |
| *RFX6* | NM_173560.4 | NC_000006.12:116895207:G:A | c.672+1G>A | – | splice donor | Pathogenic |
| *RFX6* | NM_173560.4 | NC_000006.12:116895208:T: | c.672+2del | – | splice donor | Pathogenic |
| *RFX6* | NM_173560.4 | NC_000006.12:116910932:A:G | c.673-2A>G | – | splice acceptor | Pathogenic |
| *RFX6* | NM_173560.4 | NC_000006.12:116910933:G:A | c.673-1G>A | – | splice acceptor | Pathogenic |
| *RFX6* | NM_173560.4 | NC_000006.12:116916006:G:A | c.781-1G>A | – | splice acceptor | Pathogenic |

| | | | | | | |
|---|---|---|---|---|---|---|
| *RFX6* | NM_173560.4 | NC_000006.12:116916213:T:A | c.872T>A | p.Leu291Ter | stop gained | Pathogenic |
| *RFX6* | NM_173560.4 | NC_000006.12:116916217:AC: | c.878_879del | p.His293Leufs | frameshift | Pathogenic |
| *RFX6* | NM_173560.4 | NC_000006.12:116916287:TG: | c.949_950del | p.Val317Leufs | frameshift | Pathogenic |
| *RFX6* | NM_173560.4 | NC_000006.12:116919150:TAAGAAATTTTGC: | c.1040_1052del | p.Arg347Lysfs | frameshift | Pathogenic |
| *RFX6* | NM_173560.4 | NC_000006.12:116919171:G:A | c.1058G>A | p.Trp353Ter | stop gained | Pathogenic |
| *RFX6* | NM_173560.4 | NC_000006.12:116919215:CTAA: | c.1104_1107del | p.Asp370Argfs | frameshift | Pathogenic |
| *RFX6* | NM_173560.4 | NC_000006.12:116919242:C:T | c.1129C>T | p.Arg377Ter | stop gained | Pathogenic |
| *RFX6* | NM_173560.4 | NC_000006.12:116919266:C:T | c.1153C>T | p.Arg385Ter | stop gained | Pathogenic |
| *RFX6* | NM_173560.4 | NC_000006.12:116920446:C:G | c.1320C>G | p.Tyr440Ter | stop gained | Pathogenic |
| *RFX6* | NM_173560.4 | NC_000006.12:116920454:G:A | c.1327+1G>A | – | splice donor | Pathogenic |
| *RFX6* | NM_173560.4 | NC_000006.12:116922040::GAT | c.1329_1331dup | – | splice acceptor | Pathogenic |
| *RFX6* | NM_173560.4 | NC_000006.12:116922040:G:T | c.1328-1G>T | – | splice acceptor | Pathogenic |
| *RFX6* | NM_173560.4 | NC_000006.12:116922055:G: | c.1342del | p.Val448Cysfs | frameshift | Pathogenic |
| *RFX6* | NM_173560.4 | NC_000006.12:116922061:C:T | c.1348C>T | p.Gln450Ter | stop gained | Pathogenic |
| *RFX6* | NM_173560.4 | NC_000006.12:116923111::CAAA | c.1447_1450dup | p.Asn484Thrfs | frameshift | Pathogenic |
| *RFX6* | NM_173560.4 | NC_000006.12:116923115:C:T | c.1447C>T | p.Gln483Ter | stop gained | Pathogenic |
| *RFX6* | NM_173560.4 | NC_000006.12:116923168::T | c.1506dup | p.Gly503Trpfs | frameshift | Pathogenic |
| *RFX6* | NM_173560.4 | NC_000006.12:116923181:C:T | c.1513C>T | p.Arg505Ter | stop gained | Pathogenic |
| *RFX6* | NM_173560.4 | NC_000006.12:116923224:G:T | c.1555+1G>T | – | splice donor | Pathogenic |
| *RFX6* | NM_173560.4 | NC_000006.12:116927094:C:T | c.1954C>T | p.Arg652Ter | stop gained | Pathogenic |
| *RFX6* | NM_173560.4 | NC_000006.12:116927183:C:A | c.2043C>A | p.Tyr681Ter | stop gained | Pathogenic |
| *RFX6* | NM_173560.4 | NC_000006.12:116927250:C:T | c.2110C>T | p.Gln704Ter | stop gained | Pathogenic |

| | | | | | | |
|---|---|---|---|---|---|---|
| *RFX6* | NM_173560.4 | NC_000006.12:116927268:C:T | c.2128C>T | p.Gln710Ter | stop gained | Pathogenic |
| *RFX6* | NM_173560.4 | NC_000006.12:116927295:CC:C | c.2156del | p.Pro719fs | frameshift | Pathogenic |
| *RFX6* | NM_173560.4 | NC_000006.12:116927316:C:T | c.2176C>T | p.Arg726Ter | stop gained | Pathogenic |
| *RFX6* | NM_173560.4 | NC_000006.12:116927539:G:T | c.2398+1G>T | – | splice donor | Pathogenic |
| *RFX6* | NM_173560.4 | NC_000006.12:116928756:A:G | c.2399-2A>G | – | splice acceptor | Pathogenic |
| *RFX6* | NM_173560.4 | NC_000006.12:116928810:G: | c.2452del | p.Val818Ter | frameshift | Pathogenic |
| *RFX6* | NM_173560.4 | NC_000006.12:116928815::ATCAG CACGTTTCTGTC | c.2463_2479dup | p.Ser827Thrfs | frameshift | Pathogenic |
| *RFX6* | NM_173560.4 | NC_000006.12:116928823::A | c.2463_2464ins A | p.Val822Serfs | frameshift | Pathogenic |
| *RFX6* | NM_173560.4 | NC_000006.12:116928853:CCCCC C:CCCCC | c.2499del | p.Tyr834fs | frameshift | Pathogenic |
| *RFX6* | NM_173560.4 | NC_000006.12:116928859:T: | c.2500del | p.Tyr834Thrfs | frameshift | Pathogenic |
| *RFX6* | NM_173560.4 | NC_000006.12:116882402:C:T | c.541C>T | p.Arg181Trp | missense | Likely pathogenic |

Variants listed in this table were classified as pathogenic or likely pathogenic (P/LP) according to the ACMG/AMP guidelines. Participants carrying a variant listed in this table were classified as P/LP MODY gene variant carriers. Phenotypic information from *All of Us* participants was not used for variant classification. Genomic coordinates are based on GRCh38/hg38. Sequence variants are represented in NCBI Canonical SPDI notation when available; non-SPDI representations are used for 17q12 deletions and collapsed local complex indels. cDNA and protein annotations are shown relative to the indicated RefSeq transcript. Carrier counts are not shown in accordance with the *All of Us* Data and Statistics Dissemination Policy.

**Supplementary Table 3**. Carrier prevalence and distinct pathogenic or likely pathogenic variant counts in MODY genes by sex and genetic ancestry.

| Group | Category | No. of participants | No. of distinct P/LP variants | No. of carriers | Carrier prevalence, % (95% CI) | One in *N* | *P* value |
|---|---|---|---|---|---|---|---|
| Overall | Overall | 374,973 | 203 | 370 | 0.099 (0.089 to 0.109) | 1 in 1,013 | – |
| Sex | Female | 225,728 | 136 | 217 | 0.096 (0.084 to 0.110) | 1 in 1,040 | 0.56 |
| | Male | 149,245 | 106 | 153 | 0.103 (0.087 to 0.120) | 1 in 975 | |
| Ancestry group | African | 72,346 | 54 | 76 | 0.105 (0.083 to 0.131) | 1 in 952 | 0.0034 |
| | Admixed American | 69,006 | 32 | 43 | 0.062 (0.045 to 0.084) | 1 in 1,605 | |
| | European | 217,586 | 143 | 228 | 0.105 (0.092 to 0.119) | 1 in 954 | |
| | Other ancestry | 16,035 | 18 | 23 | 0.143 (0.091 to 0.215) | 1 in 697 | |

Carrier prevalence was calculated as the number of participants carrying a pathogenic or likely pathogenic (P/LP) variant in any of the 10 MODY genes evaluated (*ABCC8*, *GCK*, *HNF1A*, *HNF1B*, *HNF4A*, *INS*, *KCNJ11*, *NEUROD1*, *PDX1*, *RFX6*) divided by the number of participants in each group. Confidence intervals (CIs) were calculated using the exact binomial method. *P* values were calculated using Fisher's exact test for sex and a chi-square test for heterogeneity across ancestry groups. Ancestry was based on *All of Us* genetic ancestry assignments and reported as African, Admixed American, European, or other ancestry. South Asian, East Asian, and Middle Eastern participants were combined as other ancestry to avoid reporting cells with fewer than 20 participants.

**Supplementary Table 4**. Distribution of MODY gene variant carriers and distinct pathogenic or likely pathogenic variants by gene.

| **Gene or gene group** | **No. of distinct P/LP variants** | **No. of carriers** | **Proportion among carriers, % (95% CI)** |
|---|---|---|---|
| *GCK* | 68 | 102 | 27.6 (23.1 to 32.4) |
| *RFX6* | 51 | 89 | 24.1 (19.8 to 28.7) |
| *HNF1A* | 38 | 73 | 19.7 (15.8 to 24.2) |
| *HNF4A* | 17 | 40 | 10.8 (7.8 to 14.4) |
| *HNF1B* | 14 | 27 | 7.3 (4.9 to 10.4) |
| Other five genes | 15 | 39 | 10.5 (7.6 to 14.1) |

Distinct pathogenic or likely pathogenic (P/LP) variants were counted as listed in **Supplementary Table 2**; the recurrent 17q12 deletion involving *HNF1B* was counted as one variant. Proportions were calculated among 370 participants carrying a P/LP variant in any of the 10 MODY genes evaluated (*ABCC8*, *GCK*, *HNF1A*, *HNF1B*, *HNF4A*, *INS*, *KCNJ11*, *NEUROD1*, *PDX1*, *RFX6*). Confidence intervals (CIs) were calculated using the exact binomial method. The “Other five genes” category includes *ABCC8*, *INS*, *KCNJ11*, *NEUROD1*, and *PDX1*, which were combined to avoid reporting cells with fewer than 20 participants.

**Supplementary Table 5.** Kaplan-Meier-estimated diabetes penetrance under primary and alternative diabetes definitions.

| Group | Definition | No. of participants | By age 40, % (95% CI) | By age 60, % (95% CI) |
|---|---|---|---|---|
| Carriers (any MODY gene) | Primary | 370 | 13.4 (10.2 to 17.6) | 43.5 (37.7 to 49.7) |
| | Medication-excluded | 370 | 12.6 (9.4 to 16.6) | 40.7 (35.0 to 46.9) |
| | Broader diabetes-label | 370 | 14.9 (11.5 to 19.2) | 45.3 (39.5 to 51.5) |
| Carriers (*GCK*) | Primary | 102 | Not shown | 56.0 (45.6 to 66.9) |
| | Medication-excluded | 102 | | 51.6 (41.1 to 62.9) |
| | Broader diabetes-label | 102 | | 58.5 (48.2 to 69.2) |
| Carriers (HNF genes) | Primary | 140 | | 45.4 (36.3 to 55.7) |
| | Medication-excluded | 140 | | 41.8 (32.8 to 52.2) |
| | Broader diabetes-label | 140 | | 48.0 (38.8 to 58.1) |
| Carriers (Non-GCK/HNF genes) | Primary | 128 | | 29.0 (20.5 to 40.2) |
| | Medication-excluded | 128 | | 29.0 (20.5 to 40.2) |
| | Broader diabetes-label | 128 | | 29.0 (20.4 to 40.1) |
| Non-carriers with top 10% T2D PRS | Primary | 37,461 | 7.1 (6.8 to 7.4) | 35.8 (35.2 to 36.4) |
| | Medication-excluded | 37,461 | 5.9 (5.7 to 6.2) | 33.2 (32.6 to 33.9) |
| | Broader diabetes-label | 37,461 | 8.0 (7.7 to 8.3) | 37.4 (36.7 to 38.0) |
| Non-carriers with middle 80% T2D PRS | Primary | 299,681 | 3.7 (3.6 to 3.8) | 18.9 (18.8 to 19.1) |
| | Medication-excluded | 299,681 | 2.6 (2.5 to 2.6) | 15.9 (15.7 to 16.1) |
| | Broader diabetes-label | 299,681 | 4.5 (4.4 to 4.6) | 20.8 (20.6 to 21.0) |
| Non-carriers with bottom 10% T2D PRS | Primary | 37,461 | 1.9 (1.7 to 2.0) | 8.6 (8.2 to 8.9) |
| | Medication-excluded | 37,461 | 0.9 (0.8 to 1.0) | 5.5 (5.2 to 5.8) |
| | Broader diabetes-label | 37,461 | 2.3 (2.1 to 2.4) | 9.8 (9.4 to 10.2) |

Diabetes penetrance was estimated using Kaplan-Meier methods with age as the time scale for the primary and two alternative diabetes definitions. Values are percentages with 95% confidence intervals (CIs). Non-carriers were stratified according to the type 2 diabetes polygenic risk score (T2D PRS) distribution among non-carriers. For the three carrier gene groups, penetrance estimates by age 40 were not reported because they were based on sparse events, in accordance with the *All of Us* Data and Statistics Dissemination Policy. HNF genes include *HNF1A*, *HNF1B*, and *HNF4A*; non-GCK/HNF genes include *ABCC8*, *INS*, *KCNJ11*, *NEUROD1*, *PDX1*, and *RFX6*.

**Supplementary Table 6**. Proportion of MODY gene variant carriers among participants with diabetes by ages 40 and 60.

| Definition | Age threshold | No. with diabetes | No. carrying a P/LP MODY gene variant | Carrier proportion, % (95% CI) |
|---|---|---|---|---|
| Primary | By age 40 | 12,592 | 45 | 0.357 (0.261 to 0.478) |
| | By age 60 | 48,152 | 122 | 0.253 (0.210 to 0.302) |
| Medication-excluded | By age 40 | 8,900 | 42 | 0.472 (0.340 to 0.637) |
| | By age 60 | 39,563 | 113 | 0.286 (0.235 to 0.343) |
| Broader diabetes-label | By age 40 | 15,120 | 50 | 0.331 (0.246 to 0.436) |
| | By age 60 | 53,370 | 129 | 0.242 (0.202 to 0.287) |

The denominator was the number of participants with diabetes at or before each age threshold, and the numerator was the number among them who carried a pathogenic or likely pathogenic (P/LP) variant in any evaluated MODY gene.

**Supplementary Table 7**. HbA1c profiles among MODY gene variant carriers and non-carriers stratified by type 2 diabetes polygenic risk score.

| HbA1c metric | Group | *N* | Median [IQR] | Adjusted difference vs. non-carriers with middle 80% T2D PRS (95% CI) | *P* value |
|---|---|---|---|---|---|
| Median HbA1c, % | *GCK* | 37 | 6.6 [6.2, 6.8] | 0.55 (0.24 to 0.86) | $4.86 \times 10^{-4}$ |
| | HNF genes | 41 | 6.8 [5.9, 7.6] | 0.68 (0.27 to 1.09) | 0.001 |
| | Non-GCK/HNF genes | 22 | 6.6 [6.0, 7.3] | 0.74 (0.09 to 1.40) | 0.027 |
| | Non-carriers with top 10% T2D PRS | 8,712 | 6.3 [5.7, 7.5] | 0.53 (0.49 to 0.57) | $2.27 \times 10^{-137}$ |
| | Non-carriers with middle 80% T2D PRS | 56,822 | 5.8 [5.5, 6.5] | Reference | – |
| | Non-carriers with bottom 10% T2D PRS | 5,746 | 5.5 [5.3, 5.8] | −0.43 (−0.45 to −0.40) | $1.30 \times 10^{-229}$ |
| Maximum HbA1c, % | *GCK* | 37 | 6.9 [6.5, 7.3] | 0.04 (−0.29 to 0.36) | 0.818 |
| | HNF genes | 41 | 7.7 [6.3, 11.3] | 1.68 (0.74 to 2.63) | $4.95 \times 10^{-4}$ |
| | Non-GCK/HNF genes | 22 | 7.5 [6.2, 9.3] | 1.08 (0.03 to 2.13) | 0.044 |
| | Non-carriers with top 10% T2D PRS | 8,712 | 7.1 [6.0, 10.5] | 0.97 (0.90 to 1.04) | $5.34 \times 10^{-145}$ |
| | Non-carriers with middle 80% T2D PRS | 56,822 | 6.1 [5.7, 7.9] | Reference | – |
| | Non-carriers with bottom 10% T2D PRS | 5,746 | 5.8 [5.5, 6.2] | −0.78 (−0.83 to −0.74) | $2.82 \times 10^{-222}$ |
| HbA1c SD, % | *GCK* | 37 | 0.25 [0.16, 0.31] | −0.16 (−0.28 to −0.03) | 0.016 |
| | HNF genes | 41 | 0.59 [0.28, 1.20] | 0.37 (0.09 to 0.66) | 0.01 |
| | Non-GCK/HNF genes | 22 | 0.42 [0.25, 0.99] | 0.16 (−0.11 to 0.42) | 0.244 |
| | Non-carriers with top 10% T2D PRS | 8,712 | 0.46 [0.22, 1.22] | 0.22 (0.20 to 0.24) | $2.01 \times 10^{-91}$ |
| | Non-carriers with middle 80% T2D PRS | 56,822 | 0.26 [0.16, 0.61] | Reference | – |
| | Non-carriers with bottom 10% T2D PRS | 5,746 | 0.20 [0.14, 0.30] | −0.17 (−0.18 to −0.16) | $1.87 \times 10^{-140}$ |

Values are shown as median [IQR]. Analyses were restricted to participants with at least three HbA1c measurements at least 90 days apart. Non-carriers were stratified into the top 10%, middle 80%, and bottom 10% of the type 2 diabetes polygenic risk score (T2D PRS) distribution among non-carriers. Adjusted differences were calculated using linear regression models with non-carriers with middle 80% T2D PRS as the reference and are reported in percentage points. Models were adjusted for age at the midpoint of the HbA1c observation window, sex, and the first 10 genetic principal components. Robust HC3 standard errors were used. HNF genes include *HNF1A*, *HNF1B*, and *HNF4A*; non-GCK/HNF genes include *ABCC8*, *INS*, *KCNJ11*, *NEUROD1*, *PDX1*, and *RFX6*.

**Supplementary Note 1**. Variant classification.

Candidate variants were selected from the *All of Us* Variant Annotation Table (VAT) among variants that overlapped coding exons or canonical splice donor or acceptor sites of the 10 MODY genes evaluated in this study (1). The initial candidate set included two variant categories: predicted loss-of-function (LoF) variants, defined as stop-gained, frameshift, or canonical splice donor/acceptor variants with LOFTEE high-confidence annotations (2), and ultra-rare missense variants with a maximum gnomAD population allele frequency <0.001% based on gnomAD allele-frequency fields in the VAT. Variants retained after this screen were reviewed according to the ACMG/AMP guidelines (3), with reference to ACGS 2020 guidance (4), applicable ClinGen guidance for variant interpretation (5, 6), and available variant-specific and gene-specific evidence. Phenotypes of *All of Us* participants were not used for variant classification. Carrier status was assigned only to participants carrying variants classified as pathogenic or likely pathogenic (P/LP) after manual review.

***Predicted loss-of-function variants***

Predicted LoF variants were not assumed to be pathogenic solely based on LOFTEE high-confidence annotation. Classification considered the expected molecular consequence in each gene, including transcript context, predicted nonsense-mediated decay (NMD), expected protein disruption, predicted splice effect when applicable, and whether heterozygous loss of function is an established disease mechanism for that gene (7). The gene-specific approach used to interpret predicted LoF variants is described below.

***ABCC8* and *KCNJ11.*** Heterozygous truncating loss of function is not established as a general mechanism of *ABCC8*- or *KCNJ11*-MODY. Pathogenic variants causing diabetes are generally associated with altered $K_{ATP}$-channel activity, particularly channel activation, whereas inactivating variants classically cause congenital hyperinsulinism (8). Therefore, predicted LoF variants in *ABCC8* and *KCNJ11* were classified as P/LP only when supported by variant-specific evidence for a MODY-related disease mechanism.

***GCK.*** Loss of glucokinase function is an established mechanism of *GCK*-MODY (6). Predicted LoF variants in *GCK* were classified as P/LP when their predicted consequence was consistent with pathogenic loss of glucokinase function under the ACMG/AMP guidelines and applicable ClinGen Monogenic Diabetes Variant Curation Expert Panel recommendations.

***HNF1A.*** Loss of function is an established mechanism of *HNF1A*-MODY (6). Predicted LoF variants in *HNF1A* were classified as P/LP when their predicted consequence was consistent with *HNF1A*-specific ClinGen Monogenic Diabetes Variant Curation Expert Panel recommendations (6). Variants with non-classical predicted consequences, such as protein extension, were interpreted using available variant-specific evidence (9).

***HNF1B.*** Heterozygous disruption of *HNF1B*, including recurrent 17q12 deletions, is a well-recognized cause of *HNF1B*-related diabetes and renal cysts and diabetes syndrome (9, 10). Predicted LoF variants in *HNF1B* were evaluated in this context, with classification considering transcript context, predicted molecular consequence, general ClinGen PVS1 recommendations, and available supporting evidence (5, 7).

***HNF4A.*** Predicted LoF variants in *HNF4A* were interpreted using applicable ClinGen Monogenic Diabetes Variant Curation Expert Panel recommendations, which assign PVS1 strength according to variant type and truncation position (6). C-terminal stop-gained or frameshift variants requiring reduced PVS1 strength were classified as P/LP only when additional evidence supported that classification.

***INS.*** *INS* truncating variants were interpreted according to predicted NMD status and expected protein consequence, rather than as simple haploinsufficiency variants. Published gene-level and family-based evidence supports heterozygous NMD-escape *INS* LoF variants as a cause of MODY (11). Variants were classified as P/LP when the predicted protein consequence and available evidence supported this disease-relevant mechanism.

***NEUROD1.*** Because *NEUROD1* is a single-exon gene, predicted LoF variants were interpreted according to the expected effect of the altered protein rather than transcript loss. Predicted LoF variants in *NEUROD1* were classified as P/LP when they were expected to disrupt a disease-relevant protein region and were supported by available evidence (11, 12).

***PDX1.*** Rare heterozygous *PDX1* variants have been reported as low-penetrance causes of MODY, and available evidence suggests that disease-relevant LoF effects depend on the predicted protein consequence and affected functional region rather than simple haploinsufficiency alone (11, 12). Predicted LoF variants in *PDX1* were therefore evaluated according to their expected protein-level effect, including whether the altered protein was predicted to affect a functionally relevant region. Variants were classified as P/LP when the predicted consequence was consistent with a disease-relevant mechanism and supported by available evidence.

***RFX6.*** Heterozygous protein-truncating variants in *RFX6* have been associated with reduced-penetrance MODY (13). Predicted LoF variants in *RFX6* were therefore evaluated as potentially disease-relevant and classified by manual review under the ACMG/AMP guidelines.

***Missense variants***
Ultra-rare missense variants were reviewed according to the ACMG/AMP guidelines. Rarity and computational prediction alone were not considered sufficient for P/LP classification. Gene-specific disease mechanisms were considered, particularly for *GCK*, in which missense variants that reduce glucokinase activity are an established mechanism of *GCK*-MODY (6). ClinVar assertions and published evidence for the variant itself, the same amino acid change, or another disease-associated change at the same residue were evaluated as variant-specific evidence (14).

**Supplementary Note 2**. Genotype and sequencing-read quality control for sequence variants.

After variant classification, participants with pathogenic or likely pathogenic (P/LP) sequence variants were identified from the short-read whole-genome sequencing (srWGS) genotype files (15). Because carrier ascertainment in this study depended on specific rare P/LP alleles, putative carrier genotypes were evaluated using target-allele-specific genotype and sequencing-read quality control. For each variant, the allele supporting the P/LP classification was treated as the target allele. Genotype information was obtained from Variant Call Format (VCF) records (16). At multiallelic sites and for indels, carrier status was assigned only when the recorded genotype contained the same target alternate allele, not merely another alternate allele at the same genomic position.

To reduce false-positive carrier calls, genotypes containing the target allele were evaluated using genotype-level filters and sequencing-read evidence. Genotype-level filtering used standard call-level metrics, including genotype quality, read depth, and allele balance (17, 18). A genotype was retained only when the target allele was present, the site-level FILTER field and genotype-level filter tag were PASS or missing, genotype quality was at least 20, and read depth calculated from allele depth was at least 10. For heterozygous genotypes, the allele balance for each called allele was required to be at least 0.20. For homozygous alternate genotypes, the target alternate-allele fraction was required to be at least 0.80. Genotypes that did not meet these criteria were excluded.

Sequencing-read support was then reviewed for calls that passed genotype-level filtering. This read-level review was performed to reduce false-positive calls arising from sequencing, mapping, genotyping, or local-alignment artifacts (18, 19). Regional alignment files were generated from the corresponding sequencing data, and read support was assessed for the target alternate allele itself rather than for the genomic position alone. A call was retained only when the target allele was supported by usable sequencing depth of at least 10, at least three usable reads carrying the alternate allele, and an alternate-allele fraction compatible with the recorded genotype. Calls were excluded when apparent alternate-allele support was attributable to poor mapping or base quality, duplicate reads, marked strand imbalance, read-end artifacts, excessive soft clipping, nearby insertion or deletion events, or local misalignment.

After genotype-level and sequencing-read review, duplicate records arising from transcript annotation or repeated representation of the same allele were collapsed so that each participant and target allele was counted once. Ambiguous or discordant records were not counted.

**Supplementary Note 3**. Ascertainment of recurrent 17q12 deletions.

Recurrent 17q12 deletions involving *HNF1B* were evaluated separately from single-nucleotide variants and indels. Structural variants were queried from the short-read whole-genome sequencing structural-variant (SV) callset in the *All of Us* Controlled Tier Curated Data Repository, version 8. This SV callset was available for 97,061 participants and was therefore available for only a subset of the full study population.

We defined the recurrent 17q12 target region on GRCh38 as chr17:36458167–37854616. SV records overlapping this region were first reviewed in the variant-level file without participant genotypes. Candidate deletions were required to be annotated as deletions (SVTYPE=DEL), have a PASS filter status, and fully span the recurrent 17q12 target region. Carrier status was then extracted from the corresponding chromosome 17 SV genotype VCF. Participants heterozygous for a deletion spanning the recurrent 17q12 region who also met the study inclusion criteria were included as *HNF1B* variant carriers.

**Supplementary Note 4**. Diabetes ascertainment.

Diabetes ascertainment was based on diagnosis codes in the electronic health record (EHR), self-reported history, laboratory measurements, and medication records available in *All of Us*. In the primary definition, diagnosis-code and self-report records were limited to type 2 diabetes because MODY is often clinically labeled as type 2 diabetes in routine care (20). Participants with at least one qualifying record were considered to have diabetes. Age at diabetes ascertainment was defined as the age at the earliest qualifying record. The criteria for the primary, medication-excluded, and broader diabetes-label definitions are summarized in the table below.

| Data source | Codes or criteria | Primary definition | Medication-excluded definition | Broader diabetes-label definition |
|---|---|---|---|---|
| Type 2 diabetes diagnosis code in the EHR | Type 2 diabetes diagnosis records, including SNOMED code 44054006 and descendant concepts where applicable | Included as a qualifying EHR-derived diagnosis-code record | Included as a qualifying EHR-derived diagnosis-code record | Included within broader EHR-derived diabetes diagnosis-code records |
| Self-reported type 2 diabetes | Participant-reported type 2 diabetes; AoU PPI concept ID 43529932 | Included as a qualifying self-report record | Included as a qualifying self-report record | Included within broader self-reported diabetes records |
| HbA1c | HbA1c measurement records, including OMOP concept ID 4184637 and LOINC codes 4548-4, 4549-2, and 17856-6; threshold ≥6.5% | Included when the value was ≥6.5% | Included when the value was ≥6.5% | Included when the value was ≥6.5% |
| Fasting glucose | Fasting glucose measurement records, including SNOMED code 167096006 and LOINC code 1558-6; threshold ≥126 mg/dL | Included when the value was ≥126 mg/dL | Included when the value was ≥126 mg/dL | Included when the value was ≥126 mg/dL |
| Non-insulin glucose-lowering medication use | Drug exposure records for non-insulin glucose-lowering medications, identified using ATC A10B-related descendants; OMOP ancestor concept ID 21600744 | Included as a qualifying medication record | Not included | Included as a qualifying medication record |
| Diabetes diagnosis code in the EHR, not restricted to type 2 diabetes | Diabetes diagnosis records using OMOP concept ID 201820 and descendant concepts | Diagnosis-code ascertainment was restricted to type 2 diabetes records | Diagnosis-code ascertainment was restricted to type 2 diabetes records | Included as a qualifying EHR-derived diagnosis-code record |
| Self-reported diabetes not restricted to type 2 diabetes | Diabetes-related participant-reported records identified from observation fields, excluding family-history-related records | Self-report ascertainment was restricted to self-reported type 2 diabetes | Self-report ascertainment was restricted to self-reported type 2 diabetes | Included as a qualifying self-report record |

HbA1c and fasting glucose thresholds followed standard diagnostic criteria for diabetes (21). Random or otherwise unspecified glucose values ≥200 mg/dL were not used as qualifying diabetes criteria because application of this threshold requires classic symptoms of hyperglycemia or hyperglycemic crisis, which cannot be reliably determined from laboratory records alone (21). Insulin use alone was not used for diabetes ascertainment because, in EHR medication data, an isolated insulin record can reflect treatment of inpatient or acute-care hyperglycemia rather than chronic diabetes (22).

The medication-excluded definition omitted non-insulin glucose-lowering medication use because some medications in this category are used for indications other than diabetes, making an isolated medication record potentially ambiguous. The broader diabetes-label definition used diagnosis-code and self-report records without restricting the documented diabetes subtype, to account for possible documentation under other diabetes labels.

**Supplementary Note References**

14. Landrum MJ, Lee JM, Riley GR, Jang W, Rubinstein WS, Church DM, et al. ClinVar: public archive of relationships among sequence variation and human phenotype. Nucleic Acids Res. 2014;42(Database issue):D980-5.
15. All of Us Research Program Genomics I. Genomic data in the All of Us Research Program. Nature. 2024;627(8003):340-6.
16. Danecek P, Auton A, Abecasis G, Albers CA, Banks E, DePristo MA, et al. The variant call format and VCFtools. Bioinformatics. 2011;27(15):2156-8.
17. Pedersen BS, Brown JM, Dashnow H, Wallace AD, Velinder M, Tristani-Firouzi M, et al. Effective variant filtering and expected candidate variant yield in studies of rare human disease. NPJ Genom Med. 2021;6(1):60.
18. DePristo MA, Banks E, Poplin R, Garimella KV, Maguire JR, Hartl C, et al. A framework for variation discovery and genotyping using next-generation DNA sequencing data. Nat Genet. 2011;43(5):491-8.
19. Zook JM, Chapman B, Wang J, Mittelman D, Hofmann O, Hide W, et al. Integrating human sequence data sets provides a resource of benchmark SNP and indel genotype calls. Nat Biotechnol. 2014;32(3):246-51.
20. Thanabalasingham G, Pal A, Selwood MP, Dudley C, Fisher K, Bingley PJ, et al. Systematic assessment of etiology in adults with a clinical diagnosis of young-onset type 2 diabetes is a successful strategy for identifying maturity-onset diabetes of the young. Diabetes Care. 2012;35(6):1206-12.
21. American Diabetes Association Professional Practice Committee for D. 2. Diagnosis and Classification of Diabetes: Standards of Care in Diabetes-2026. Diabetes Care. 2026;49(Supplement_1):S27-S49.
22. Korytkowski MT, Muniyappa R, Antinori-Lent K, Donihi AC, Drincic AT, Hirsch IB, et al. Management of Hyperglycemia in Hospitalized Adult Patients in Non-Critical Care Settings: An Endocrine Society Clinical Practice Guideline. J Clin Endocrinol Metab. 2022;107(8):2101-28.
